# 40 Hz Light Flickering Promotes Sleep through Cortical Adenosine Signaling

**DOI:** 10.1101/2023.10.07.23296695

**Authors:** Yan He, Xuzhao Zhou, Tao Xu, Zhaofa Wu, Wei Guo, Xi Xu, Yuntao Liu, Yi Zhang, Huiping Shang, Zhimo Yao, Zewen Li, Zhihui Li, Tao Feng, Shaomin Zhang, Rodrigo A. Cunha, Zhili Huang, Yulong Li, Xiaohong Cai, Jia Qu, Jiang-Fan Chen

**Affiliations:** The Eye and Brain Center, State Key Laboratory of Ophthalmology, Optometry and Visual Science, Eye Hospital, Wenzhou Medical University, Wenzhou, 325027, China; Oujiang Laboratory (Zhejiang Laboratory for Regenerative Medicine, Vision and Brain Health), School of Ophthalmology & Optometry and Eye Hospital, Wenzhou Medical University, Wenzhou, 325027, China; Department of Pediatrics, The Second Affiliated Hospital and Yuying Children’s Hospital of Wenzhou Medical University, Wenzhou, Zhejiang 325027, China; State Key Laboratory of Membrane Biology, School of Life Sciences, Peking University, Beijing 100871, China; Institute of Genomic Medicine, Wenzhou Medical University, Wenzhou, Zhejiang Province 325035, China; Key Laboratory of Biomedical Engineering of Ministry of Education, Qiushi Academy for Advanced Studies, Zhejiang University, 38 Zheda Road, Hangzhou, 310027 China; CNC-Center for Neuroscience and Cell Biology, Faculty of Medicine, University of Coimbra, 3004-504 Coimbra, Portugal; Department of Pharmacology, School of Basic Medical Sciences; State Key Laboratory of Medical Neurobiology and MOE Frontiers Center for Brain Science, and Institutes of Brain Science, Fudan University, Shanghai, 200032, China

**Keywords:** 40 Hz light flickering, adenosine, equilibrative nucleoside transporter, sleep, visual cortex

## Abstract

Flickering light stimulation has emerged as a promising non-invasive neuromodulation strategy to alleviate neuropsychiatric disorders. However, the lack of a neurochemical underpinning has hampered its therapeutic development. Here, we demonstrate that light flickering triggered an immediate and sustained increase (up to 3 hours after flickering) in extracellular adenosine levels in the primary visual cortex and other brain regions, as a function of light frequency, intensity, and wavelength, with maximal effects observed at 40 Hz frequency. We discovered cortical (glutamatergic and GABAergic) neurons, rather than astrocytes, as the cellular source, and intracellular adenosine generation from calcium influx-triggered, AMPK-associated energy metabolism pathways (but not SAM-transmethylation or salvage purine pathways) and adenosine efflux mediated by equilibrative nucleoside transporter-2 (ENT2) as the molecular pathway responsible for extracellular adenosine generation. Importantly, 40 Hz light flickering for 30 min enhanced sleep in mice in a frequency-dependent manner. This somnogenic effect was absent in mice lacking ENT2 but replicated by administering adenosine to the visual cortex. Brief 40 Hz light flickering also promoted sleep in children with insomnia by decreasing sleep onset latency, increasing total sleep time, and reducing waking after sleep onset. Collectively, our findings establish adenosine signaling via ENT2 as the neurochemical basis for 40 Hz flickering-induced sleep and unravel a novel and non-invasive treatment for insomnia, a condition that affects 20% of the world population.

## Introduction

One of the most significant environmental factors affecting human physiology is light (*1*). As a non-invasive neuromodulation strategy, light stimulation has demonstrated potential for alleviating various pathological changes in animal models of depression (*2, 3*), insomnia (*4*), and Alzheimer’s disease (AD) (*5*). In particular, light flickering at 40 Hz is receiving increased attention for its ability to reverse pathological features of AD (*5–7*), ischemia (*8*), and traumatic brain injury (*9*) in animals. The demonstration of gamma oscillation entrainment across multiple brain regions, including the hippocampus (*6*), and the reduction of amyloid loading with activated microglia and increased vascular densities and improvement of cognitive performance by combined visual and acoustic stimulation (GENUS) in AD mice (*5, 7*), has led to the US FDA designating the 40 Hz flicker paradigm (GENUS equipment) as a “breakthrough device” in 2021 for further clinical investigation (*10*). Light flickering-induced neurochemical coupling mechanisms are the key to its therapeutic effect. Current research focuses on the electrophysiological responses induced by light, such as gamma oscillation entrainment, as a crucial factor in inducing therapeutic effects (*6, 7, 11*). However, the neurochemical and molecular adaptations that determine the biological effects of 40 Hz light flickering are still unexplored, even though the treatment often requires more than 2 weeks to 2 months to achieve effective therapeutic benefits (*5–7*). The lack of understanding of this central issue has hindered the development of effective therapies and contributed to the variable effects of 40 Hz flickering (*11*). Thus, it is paramount to delineate the neurochemical mechanisms underlying 40 Hz light flickering to fully exploit its therapeutic potential.

Light flickering-triggered neural activity (e.g., spike activity or gamma oscillations) is especially energy demanding (*12*), requiring robust mitochondrial function and producing strong oxygen consumption with hemodynamic changes, as evidenced by blood oxygen level-dependent (BOLD) signals in the cortex (*13, 14*). Given that adenosine is the main signaling molecule resulting from increased energy expenditure (*15–17*), we reasoned that light flickering- triggered neural activity represents a form of energy-demanding neural stimulation that leads to rapid adenosine generation. Extracellular adenosine, with its intrinsic link with energy metabolism, can simultaneously act as a modulator of neurotransmission and synaptic plasticity (*18*) and a homeostasis regulator of metabolism, motility, proliferation, and vasodilation (*16, 19*). Therefore, we hypothesized that 40 Hz light flickering induces brain adenosine signaling to produce therapeutic effects.

Among its multiple physiological and pathophysiological effects, adenosine is a well-known physiological regulator of homeostatic sleep needs. Extracellular adenosine levels in the basal forebrain rise after prolonged wakefulness, resulting in increased sleep pressure (*20–22*). Adenosine’s somnogenic effect can be highlighted through the intracerebral or systemic administration of adenosine and its analogs or by pharmacological modulation of adenosine metabolism (*23*). Conversely, caffeine, the most widely consumed psychostimulant, and a non-selective adenosine receptor antagonist, produces a strong arousal effect (*22–24*). Based on this reasoning, we propose that 40 Hz light flickering could potentially increase cortical adenosine levels to achieve a somnogenic effect, offering a novel and non-invasive therapy for insomnia, which affects 20% of the world population.

In this study, we established that cortical adenosine signaling is crucial for the biological effects of 40 Hz light flickering on sleep. We demonstrated here that light flickering triggered a rapid and sustained increase in extracellular adenosine levels in the visual cortex and other brain regions, which depended on the light flickering frequency (with 40 Hz flickering producing the maximal effect), intensity, and wavelength. We identified cortical (glutamatergic and GABAergic) neurons (rather than astrocytes) as the cellular source of extracellular adenosine generated by 40 Hz light flickering. Furthermore, we revealed that the calcium influx-, AMPK phosphorylation-associated intracellular adenosine generation from energy metabolism and adenosine efflux mediated by equilibrative nucleoside transporter-2 (ENT2, not ENT1) were the molecular cascade responsible for extracellular adenosine generation. Importantly, 40 Hz light flickering for 30 minutes promoted sleep onset and maintenance in mice and children with insomnia. Collectively, we have defined cortical adenosine signaling via ENT2 as the neurochemical basis for 40 Hz flickering-induced sleep, and developed a novel and non-invasive treatment for insomnia with potentially wider therapeutic implications.

## Results

### 1. 40 Hz light flickering induces an immediate and sustained increase in extracellular adenosine levels in the visual cortex

To examine the role of extracellular adenosine in the biological effects of 40 Hz light flickering, we utilized a highly sensitive and selective G protein-coupled receptor (GPCR)- activation-based (GRAB) adenosine sensor (GRAB_Ado_) to monitor changes in extracellular adenosine levels in response to 40 Hz light flickering(*21*). After injecting an AAV expressing GRAB_Ado_ or a non-binding mutant of GRAB_Ado_ into the primary visual cortex (V1) (Fig. 1A), we employed fiber photometry to assess fluorescence signals indicative of extracellular adenosine. After exposing mice to white light flickering (3000 lux, 40 Hz, 50% duty cycle) produced by a custom-made LED device (*5, 25*), we found an immediate and sustained rise in extracellular adenosine levels during and after the 30 min 40 Hz light flickering period. In addition to the immediate surge in extracellular adenosine levels, we observed a substantial and delayed increase in adenosine levels in V1 after the cessation of 40 Hz light flickering. This increase reached its peak (with ΔF/F value up to 20-25% higher than baseline) at 30 min and lasted for up to 3 h before returning to baseline levels (Fig. 1D). The specificity of this extracellular adenosine generation was validated by the absence of fluorescent signal increases in V1 neurons expressing the non-binding mutant of GRAB_Ado_ (Fig. 2A). Importantly, we found that the effect of light flickering on extracellular adenosine levels in the visual cortex was frequency-dependent: 40 Hz light flickering produced the maximum increase in extracellular adenosine levels that lasted the longest (up to 3 h), while light flickering at 20 Hz or 80 Hz resulted in discrete changes in extracellular adenosine levels in V1 (Fig. 1D). Thus, the frequency of light flickering plays a crucial role in determining the increase in extracellular adenosine levels in V1.

**Fig. 1.**
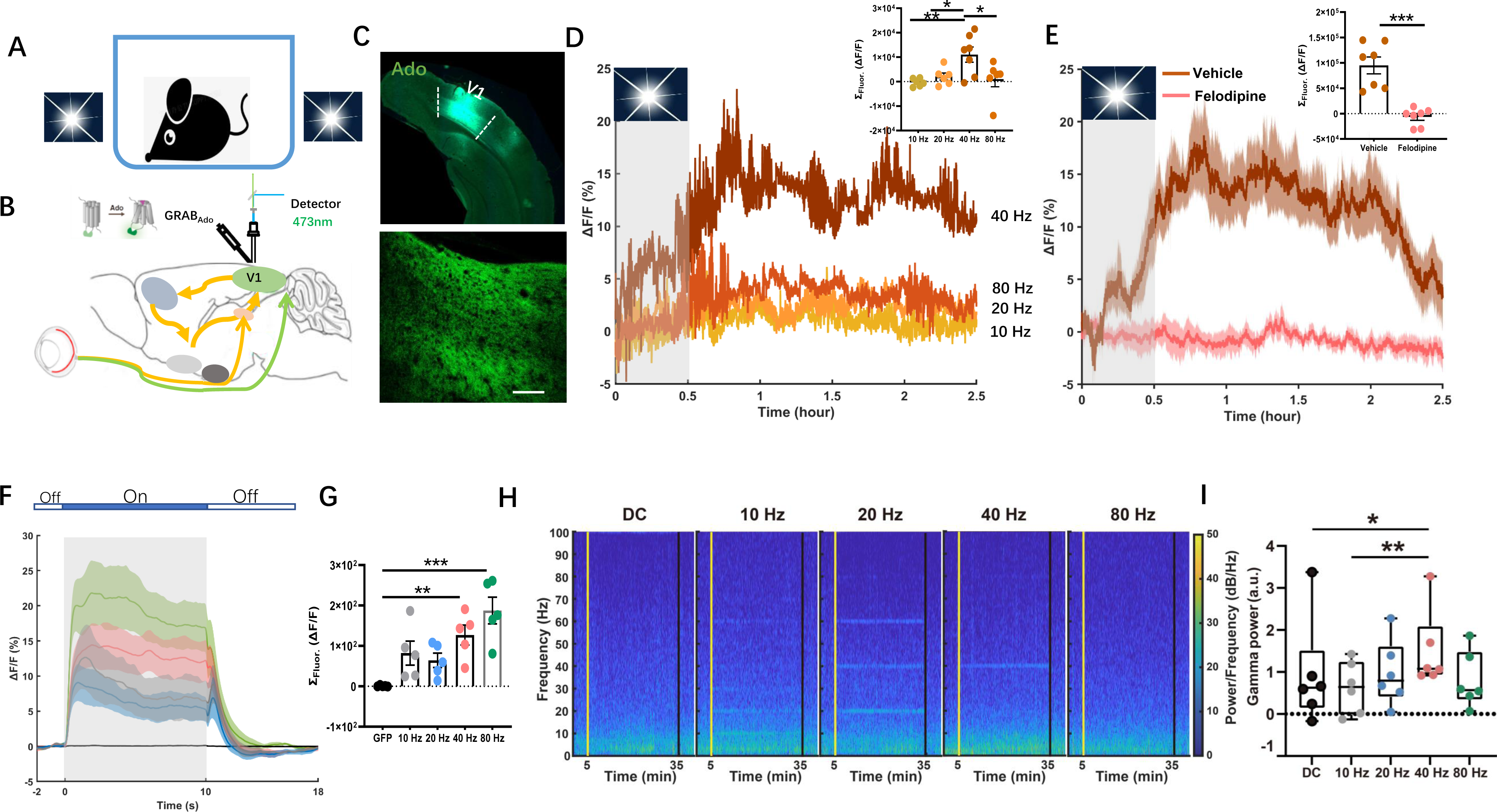
40 Hz light flickering induces a robust and sustained increase in extracellular adenosine levels in the primary visual cortex in correlation with LFP gamma power. A. Schematic diagram depicting the experimental configuration for visual stimulation. B. Schematic diagram depicting fiber photometry recording of extracellular adenosine levels in the V1 visual cortex using GRABAdo1.0. C. Fluorescence image of V1 showing the expression of hSyn-GRABAdo1.0. D. Light flickering at 10 Hz, 20 Hz, 40 Hz, and 80 Hz induced an immediate (within minutes) and sustained (lasting for up to 3 h) increase in the extracellular adenosine levels in V1 during and after flickering, with 40 Hz flickering yielding the maximum increase. Insert panel: quantification of light flickering-evoked adenosine signals (n = 5-7/group). E. Pretreatment with an L-type VGCC inhibitor felodipine (10 mg/kg, i.p.) abolished the increased extracellular adenosine levels induced by 40 Hz flashing (n = 7/group). F-G. Light flickering at 10 Hz, 20 Hz, 40 Hz, and 80 Hz (50% duty cycle) produced a linear rise in calcium signals in V1 when comparing the “On time” (10 s) with the “Off time”, with 80 Hz flickering producing the strongest amplitude of the calcium peak. H. Representative time-resolved spectrograms of the primary visual cortex (V1) during 10Hz, 20 Hz, 40 Hz, and 80 Hz light flickering and DC stimulation. The vertical line in each plot represents the start and end of the light-flickering period. I. Quantified and normalized values of gamma power during flickering (n = 6/group). The skewed data were analyzed using repeated measured Friedman test followed by Dunn’s multiple comparisons test. The data are presented as mean ± SEM or upper and lower quartile (IQR) and median (boxplot), ***P < 0.001, **P < 0.01, *P < 0.05.

**Fig. 2.**
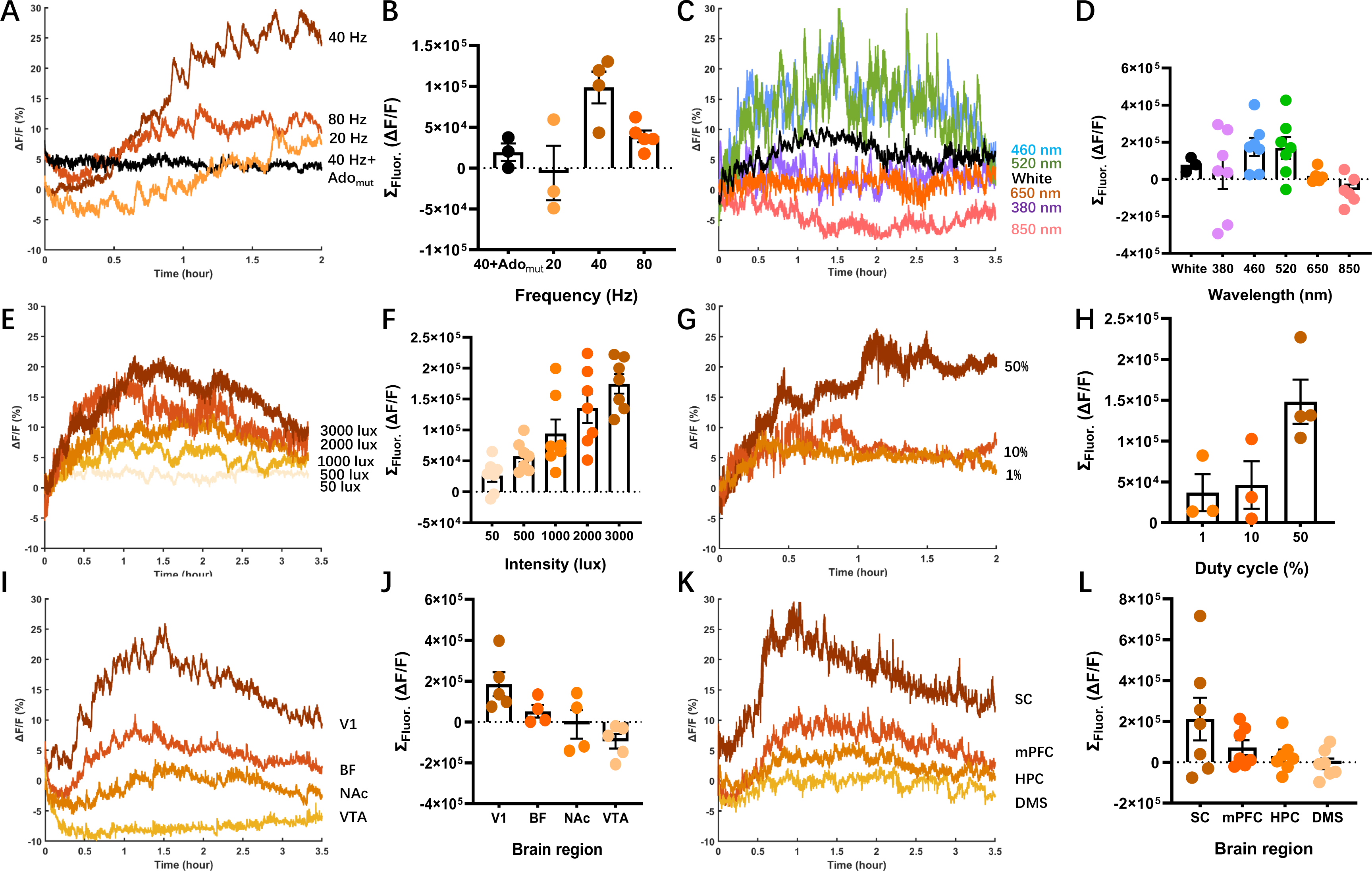
Light flickering increases extracellular adenosine levels in light frequency-, wavelength-, intensity-, and duty cycle-dependent manners in V1 and other brain regions. A-B. Extracellular adenosine levels increase specifically in response to 40 Hz light flickering compared to 20 Hz and 80 Hz light flickering (n = 3∼5/group). C-D. The effect of 40 Hz light flashing at different wavelengths (500 lux) on extracellular adenosine levels in V1 visual cortex (White light = 3, 380- 850 nm wavelengths n = 7/group). E-F. The effect of 40 Hz white light flickering with different intensities (50-3000 lux) on extracellular adenosine levels in V1 (n = 7/group). G-H. The effect of 40 Hz light flashing at 3000 lux intensity with varying duty cycles (1%, 10%, and 50% on time) on extracellular adenosine levels (n = 3∼4/group). I-L. The effect of 40 Hz light flickering (3000 lux) on extracellular adenosine levels in eight brain regions of mice, including V1, basal forebrain (BF), nucleus accumbens (NAc), ventral tegmental area (VTA), superior colliculus (SC), medial prefrontal cortex (mPFC), hippocampus (HPC) and dorsal medial striatum (DMS) (n= 4/group). The data were presented as mean ± SEM.

We addressed the role of calcium signaling in light flickering-induced adenosine release, as we recently reported that L-type voltage-gated calcium channels (L-VGCC) are essential for adenosine release induced by high-frequency electric stimulation (*26*). We found that a VGCC blocker, felodipine (10 mg/kg, i.p.) almost completely abolished the 40 Hz flickering-induced increase in extracellular adenosine levels compared to the vehicle group (Fig. 1E). The effects of an another L-VGCC nimodipine (10 mg/kg, i.p.) were similar (Extended Data Figs. 1), consistent with a recent study that RGS12-regulated VGCC mediate the 30-50 Hz light flickering effect on synaptic plasticity in hippocampus (*8*). Next, we examined the effect of light flashing at different frequencies (10-80 Hz) on calcium signaling in V1 using fiber optometry and we observed a linear increase in calcium signals with increasing frequency. Specifically, light flickering at 10 Hz, 20 Hz, 40 Hz, and 80 Hz resulted in a linear rise in calcium signals in V1, with the strongest response observed at 80 Hz, producing the highest amplitude of the calcium peak (Fig. 1F and 1G). These results suggest that while calcium influx is required for the induction of adenosine release, it alone cannot explain the frequency-specificity of light flickering.

We then aimed to establish a correlation between extracellular adenosine generation and gamma oscillation power in response to light flickering at different frequencies by recording local field potentials (LFPs) before, during, and after the light flickering phase (30 min). We found that the LFP oscillation was frequency-dependent in response to light flickering (3000 lux), where each frequency of light flickering elicited a specific LFP oscillation at that frequency, as well as at its higher harmonic frequencies (Fig. 1H and Extended Data Figs. 2A to E). We compared the LFP power within the flickering frequency and found that the normalized power significantly increased at both 20 Hz and 40 Hz compared to direct current (DC) (20 Hz *vs.* DC, P = 0.0041; 40 Hz *vs.* DC, P = 0.0005) (Extended Data Figs. 2F). Since gamma oscillation has been shown to be strongly correlated with a hemodynamic response (i.e., energy demand) (*13*), we conducted a power analysis of LFP oscillations specifically within the gamma frequency range (22-90 Hz) (*13*). We discovered that among all flickering frequencies, 40 Hz produced the strongest gamma power (40 Hz *vs.* 10 Hz, P = 0.0076; 40 Hz *vs.* DC, P = 0.0423) (Fig. 1I), suggesting that 40 Hz light flickering has the highest energy demand compared to other frequencies.

## 2. Light flickering increases extracellular adenosine levels in frequency-, wavelength-, intensity-, and duty cycle-dependent manners in V1 and other brain regions

Next, we characterized the effect of different light flickering parameters (namely frequency, intensity, wavelength, and duty cycle stimuli) on the induction of extracellular adenosine levels in V1 and other brain regions. First, as shown in Fig. 2, we confirmed the frequency-specific effect of light flickering on the sustained elevation of extracellular adenosine levels in V1: the sustained increase in extracellular adenosine levels in response to light flickering was specific to 40 Hz, as minimal or no changes were observed in response to 20 Hz or 80 Hz light flickering (Fig. 2A and B). Moreover, frequency-dependent of light flickering on the extracellular adenosine levels was also observed in the basal forebrain, with 40 Hz flickering producing higher extracellular adenosine than 20 Hz flickering Extended Data Figs. 1).

Second, we determined the effect of 40 Hz flickering light with different wavelengths on extracellular adenosine production in V1 (Fig. 2C and D). We observed that 40 Hz flickering (500 lux) at 460 nm (green) and 520 nm (blue) significantly raised extracellular adenosine levels; the time course with the different wavelengths was generally similar, with a delayed increase starting at 10 min, reaching a peak at 30 min, and lasting for 2-3 h. Notably, the levels of extracellular adenosine elicited by 40 Hz flashing at 460 nm and 520 nm were higher than those produced by white light (500 lux). In contrast, 40 Hz flickering at wavelengths of 380 nm, 650 nm, and 850 nm failed to elevate extracellular adenosine levels (Fig. 2C and D). Thus, the light wavelength has a substantial impact on 40 Hz flickering effects on extracellular adenosine generation.

Third, we investigated the impact of varying intensities of white light (50, 500, 1000, 2000, and 3000 lux) on extracellular adenosine production in the visual cortex in response to 40 Hz flickering. We found that 40 Hz flickering at high light intensities (2000 and 3000 lux) induced significantly higher levels of extracellular adenosine in V1, with a peak ΔF/F of 22%. This effect persisted for a longer duration (2-3 h) compared to the effect induced by 40 Hz flickering at low intensities (50 and 500 lux), as shown in Fig. 2E and F. Importantly, the amplitude of the rise in extracellular adenosine levels depended on the intensity of 40 Hz light flickering. We also investigated how different duty cycles (i.e., the duration of the on-time during the cycle) of 40 Hz light flickering stimulation affected extracellular adenosine levels in V1 (Fig. 2G). Increasing the duty cycle, which results in higher light exposure intensity, produced a greater increase in extracellular adenosine levels. Specifically, 40 Hz flickering with a 50% duty cycle (i.e., 12.5 ms light-on and 12.5 ms light-off) produced a greater rise in extracellular adenosine than 40 Hz flashing with 10% or 1% duty cycles, further supporting the critical role of light intensity (Fig. 2H). These findings indicate that light intensity plays a crucial role in inducing extracellular adenosine in response to 40 Hz light flickering.

Finally, we inserted a multi-fiber probe into the brains of mice at eight different brain regions to record the regional variations in extracellular adenosine levels induced by 40 Hz light flickering (Fig. 2I to L). There was a robust increase in extracellular adenosine levels in V1 and the superior colliculus (SC), with a ΔF/F of 20-30% above the baseline, followed by a moderate increase in the basal forebrain (ΔF/F of 10-15%), prefrontal cortex (ΔF/F of 5-10%), and hippocampus (ΔF/F of 0-5%), as depicted in Fig. 2I to L. The temporal features of adenosine elevation induced by 40 Hz flickering were similar across these brain regions, with little to no increase observed in the striatum and ventral tegmental area (Fig. 2I to L).

### 3. Glutamatergic and GABAergic neurons are the main cellular source of light flickering-induced adenosine generation in V1

As adenosine can be released from both astrocytes and neurons (*17, 27*), we further identified the cellular source responsible for the extracellular adenosine generation induced by 40 Hz light flickering in V1. We first assessed the involvement of astrocytes in extracellular adenosine production induced by 40 Hz flickering by inhibiting astrocyte Ca^2+^ signaling using hPMCA2w/b expression (Fig. 3A to C). We confirmed the selective expression of hPMCA2w/b in V1 astrocytes by immunohistochemistry, which showed its co-localization with GFAP immunoreactivity (Fig. 3B). The foot shock conditioning tasks revealed that expression of AAV2/9-GfaABC1D-hPMCA2w/b in astrocytes partially suppressed astrocytic activation, as evidenced by a reduction in the Ca^2+^ signal response in V1 compared to mice expressing AAV2/9-GfaABC1D-mCherry, which is consistent with a previous study (*28*) (Fig. 3C). Next, we co-microinjected AAV2/9-GfaABC1D-hPMCA2w/b and GRAB_Ado_ into a single hemisphere of the visual cortex, while AAV2/9-GfaABC1D-mCherry and GRAB_Ado_ were injected into the other hemisphere. Two weeks after the injection, the increase in extracellular adenosine induced by 40 Hz flickering in V1 was not affected by the partial inhibition of astrocytic activity caused by hPMCA2w/b compared to the mCherry group (Fig. 3D), indicating that astrocytic activity does not play a crucial role in extracellular adenosine generation induced by light flickering. This result is consistent with a recent study showing that electric stimulation fails to induce adenosine release from astrocytes (*26*).

**Fig. 3.**
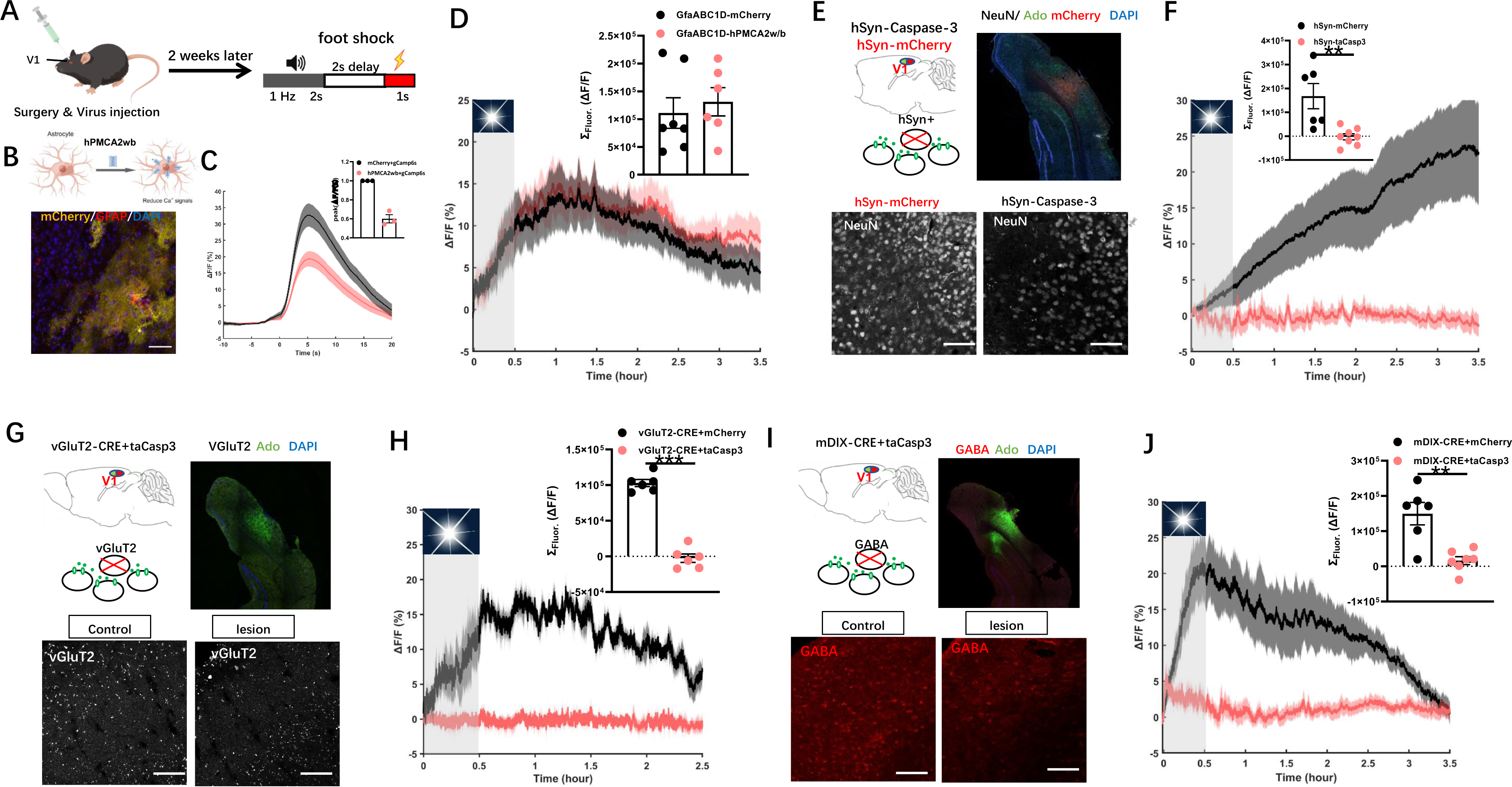
Glutamatergic and GABAergic neurons are the primary cellular source for 40 Hz flickering-induced extracellular adenosine generation in V1. A. Behavioral protocols for reward and foot shock conditioning tasks. B. Immunohistochemical (IHC) images showing the co- localization of GFAP and hPMCA2w/b in V1 astrocytes. C. Expression of hPMCA2w/b in V1 partially reduced astrocytic activity, as indicated by calcium signaling using GCaMP detector during foot shock conditioning tasks (n = 3/group). D. Partial inhibition of astrocytic activity by hPMCA2w/b expression had no effect on extracellular adenosine generation in response to 40 Hz flickering (n = 6-7/group). E. Schematic diagram depicting the fiber photometry recording of extracellular adenosine levels after selective ablation of total neurons in V1 using Caspase-3 (upper left panel); fluorescence image of V1 showing the staining of NeuN in V1 carrying AAV2/9-EF1a- flex-taCasp3 or AAV2/9-EF1a-flex-mCherry. F. Ablation of V1 neurons significantly reduced the generation of extracellular adenosine in response to 40 Hz light flickering compared to control mice (n = 6-7/group). G. Schematic diagram depicting the strategy used to selectively ablate vGluT2+ neurons in the visual cortex using vGluT2-CRE mice coupled with AAV2/9-EF1a-flex-taCasp3 (upper left panel). Fluorescence image of V1 showing the reduction of vGluT2+ neurons in V1 expressing AAV2/9-EF1a-flex-taCasp3 compared to the cortex expressing AAV2/9-EF1a-flex-mCherry (lower panel). H. Ablation of vGluT2+ neurons in V1 suppressed the generation of extracellular adenosine in response to 40 Hz flickering compared to control mice (n = 6/group). I. Schematic diagram depicting the strategy used to selectively ablate GABA neurons in V1 using AAV2/9-mDIX-CRE coupled with AAV2/9-EF1a-flex-taCasp3 (upper left panel). Fluorescence image of V1 showing the reduction of GAD+ neurons in V1 expressing AAV2/9-EF1a-flex-taCasp3 compared to V1 expressing AAV2/9-EF1a-flex-mCherry (lower panel). J. Ablation of the GAD+ neurons in V1 suppressed the generation of extracellular adenosine in response to 40 Hz flickering compared to control mice (n = 6-7/group). ***P < 0.001, **P < 0.01, *P < 0.05, Student’s t-test. The data are mean ± SEM.

Next, we investigated the role of cortical neurons in extracellular adenosine generation induced by 40 Hz light flickering using hSyn-Caspase-3 to induce selective neuronal apoptosis (*21*). We co-injected AAVs carrying GfaABC1D-GRAB_Ado_ and hSyn-Caspase-3 into the visual cortex of one hemisphere and GfaABC1D-GRAB_Ado_ with hSyn-mCherry into the other hemisphere. Two weeks after achieving targeted expression, we confirmed using NeuN immunoreactivity the selective reduction in the number of neurons in V1 expressing hSyn- Caspase-3 compared to the cortex expressing hSyn-mCherry (Fig. 3E). We found that in the hemisphere expressing mCherry, the extracellular adenosine levels gradually and progressively increased during 40 Hz light flickering and persisted for 180 min after the cessation of stimulation (Fig. 3F). Notably, the pattern of increased extracellular adenosine levels observed via GRAB_Ado_ expressed in astrocytes exhibited some dissimilarity compared to the pattern detected by GRAB_Ado_ expressed in neurons, indicating a subtle variation in local adenosine levels between neurons and astrocytes. Importantly, the visual cortex that expressed hSyn-Caspase-3 did not exhibit any progressive rise in extracellular adenosine levels in response to 40 Hz light flickering. This suggests that V1 neurons are the primary cellular source of extracellular adenosine generation induced by 40 Hz light flickering.

We further dissected the specific contribution of glutamatergic neurons to extracellular adenosine generation by depleting glutamatergic neurons in the visual cortex. In vGluT2-CRE mice, we co-injected AAV2/9-EF1a-flex-taCasp3 together with hSyn-GRAB_Ado_ into one hemisphere (the visual cortex) and AAV2/9-EF1a-flex-mCherry together with hSyn-GRAB_Ado_ into the other hemisphere. Two weeks after injection, we confirmed by vGluT2 immunofluorescence staining the selective reduction of vGluT2^+^ neurons in V1 expressing hSyn- Caspase-3 compared to the cortex expressing hSyn-mCherry (Fig. 3G). In response to 40 Hz light flickering, V1 neurons expressing vGluT2-mCherry exhibited a substantial increase in extracellular adenosine levels (Fig. 5H). However, the depletion of glutamatergic neurons in V1 almost completely abolished the production of extracellular adenosine triggered by 40 Hz flickering, both during the initial and delayed phases, supporting the idea that glutamatergic neurons are a primary cellular source of extracellular adenosine in response to 40 Hz light flickering.

Finally, we determined the specific contribution of GABAergic neurons to extracellular adenosine generation by depleting GABAergic neurons in V1 via co-injecting AAV2/9-EF1a- flex-taCasp3 and AAV2/9-mDIX-CRE together with hSyn-GRAB_Ado_ into one hemisphere and AAV2/9-EF1a-flex-mCherry and AAV2/9-mDIX-CRE together with hSyn-GRAB_Ado_ into the other hemisphere. Two weeks after the injections, we verified by GABA immunofluorescence staining the selective reduction of GABA neurons in the visual cortex expressing hSyn-Caspase- 3 compared to the cortex expressing hSyn-mCherry (Fig. 5I). We found that the hemisphere expressing AAV2/9-EF1a-flex-mCherry showed a robust and progressive elevation in extracellular adenosine upon exposure to 40 Hz flickering and lasted for 3 h after flickering cessation. In contrast, 40 Hz flickering-induced extracellular adenosine generation was largely attenuated in V1 expressing AAV2/9-EF1a-flex-taCasp3 (Fig. 5J). Thus, GABAergic neurons are also a cellular source of extracellular adenosine in the visual cortex in response to 40 Hz light flickering.

### 4. The ENT2-mediated efflux of intracellular adenosine is the main source for 40 Hz flickering-induced extracellular adenosine generation in V1

We next aimed to identify the pathways responsible for the 40 Hz flickering-induced increase in extracellular adenosine that occurs after flickering stimulation cessation, distinguishing between extracellular and intracellular mechanisms (Fig. 4A). For extracellular adenosine production, 40 Hz flickering may trigger ATP release from neurons and astrocytes, followed by extracellular conversion to AMP and subsequently to adenosine via ecto- nucleotidases CD39 and CD73, respectively. To address this route, we first employed an ATP sensor (GRAB_ATP1.0_)(*29*) to monitor the dynamics of extracellular ATP levels in V1 in response to 40 Hz light flickering (Fig. 4B-C). The validity of the ATP probe was confirmed by the expression of GRAB_ATP1.0_ (Fig. 4B), and ATP events were identified in V1 following the footshock task (insert figure in Fig. 4C) and LPS treatment (Extended Data Figs. 3) during a 120- min period, which is consistent with previous reports (*29*). We observed that 40 Hz flickering did not increase ATP events in V1, as detected by GRAB_ATP1.0_ (Fig. 4C). Furthermore, we observed that extracellular adenosine levels in V1 of both wild-type and *CD73*-KO mice displayed a largely similar delayed increase, peaking at 30 min and lasting for 3 h upon 40 Hz flickering stimulation (Fig. 4D). Thus, the extracellular formation of adenosine by ATP release and CD73-dependent conversion is not the primary source of the 40 Hz flickering-induced increase in extracellular adenosine in V1.

**Fig. 4.**
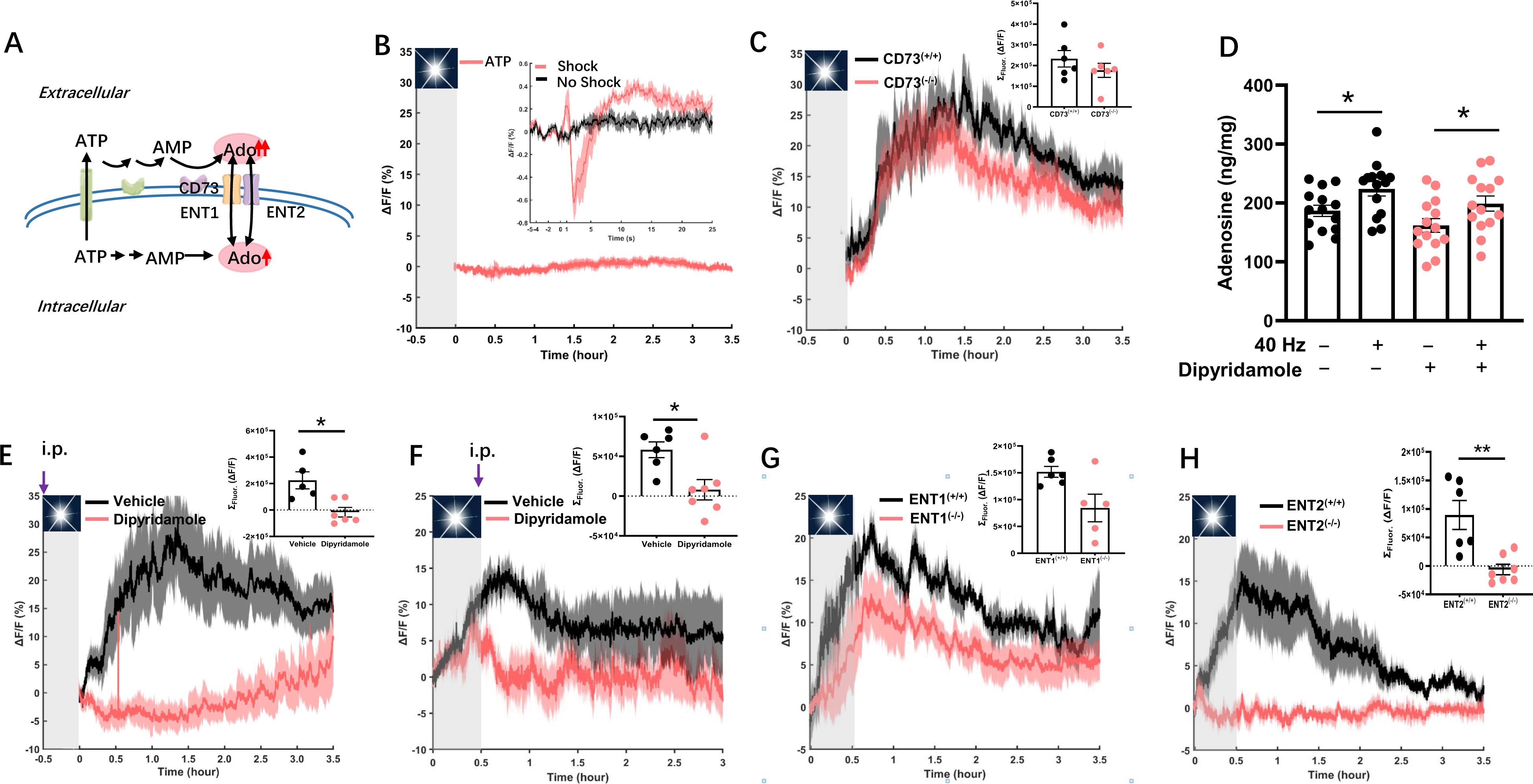
Extracellular adenosine generation in response to 40 Hz flickering in the primary visual cortex is mediated by the equilibrative nucleoside transporter 2 (ENT2). A. Schematic representation of extracellular adenosine generation through the extracellular pathway (i.e., ATP release from neurons and astrocytes, followed by extracellular conversion to AMP and adenosine via ecto-nucleotidases CD73) or intracellular pathway (i.e., as a by-product of increased energy metabolism and other pathways, followed by an ENT1/2-mediated transmembrane efflux of adenosine). B. Measurement of extracellular ATP levels after electric shock in mice expressing GRABATP (insert figure) and after 40 Hz flickering, (n = 6/group). C. Extracellular adenosine increase (as detected by GRABAdo) caused by 40 Hz light flickering in V1 of WT and CD73-KO mice with average values displayed in the insert (n = 6/group). D. Total tissue (intracellular and extracellular) adenosine levels in V1, as assessed by HPLC analysis, after 40 Hz light flickering with or without pretreatment with dipyridamole. (n = 14/group); *P < 0.05 comparing the dipyridamole-treated group with the vehicle-treated group. E. Pretreatment with dipyridamole (30 min prior to light flickering) abolished 40 Hz flickering-induced the extracellular adenosine generation during light flickering and after stimulation cessation (n = 7/group). F. Administering dipyridamole immediately after light flickering also eliminated 40 Hz flickering-induced subsequent extracellular adenosine generation after light flashing cessation (n = 6/group). G. 40 Hz flickering induced robust and sustained increase of the extracellular adenosine generation in both WT and *ENT1*-KO mice, although slightly lower than in WT mice at later time points (*ENT1*-KO: n = 5-6/group). H. 40 Hz flickering-induced extracellular adenosine generation was robust and sustained in WT but largely abolished in *ENT2*-KO mice (*ENT2*-KO: n = 6-7/group). The data are presented as mean ± SEM, **P < 0.01, *P < 0.05, Student’s t-test; WT vs. KO; dipyridamole-treated group vs. vehicle group.

**Fig. 5.**
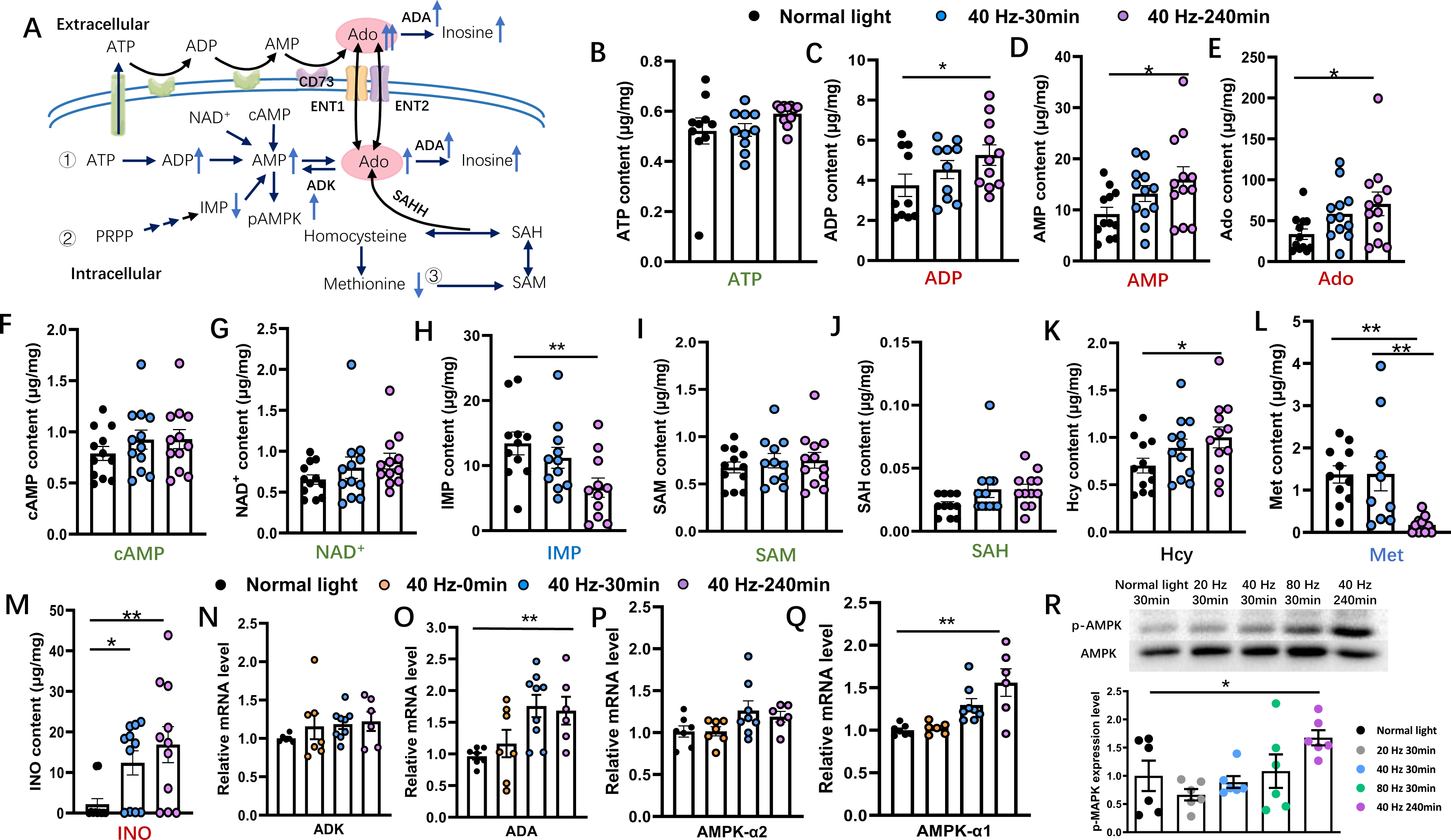
The intracellular pathway of adenosine production in response to 40 Hz flickering primarily involves energy metabolism. A, Description of the extracellular and intracellular pathways involving multiple adenosine-generating and degradation pathways. Panels B-G, present the effect of 30-min 40 Hz flickering on the intracellular metabolic changes, including major adenosine-generating, adenosine-degrading, and efflux pathways in V1 assessed 30 min after flickering cessation by the targeted UHPLC-MS/MS analysis. ATP (B), ADP (C), AMP (D), adenosine (Ado, E), cAMP (F), and NAD+ (G). *P < 0.05 for the 40 Hz-treated vs. normal light groups at 30 min; **P < 0.01 when comparing 40 Hz vs. normal light groups at 240 min. H, The effect of 40 Hz light flickering on the level of IMP, a main component in the de novo/salvage pathway, in V1. **P < 0.01 when comparing 40 Hz vs. normal light groups at 240 min. I-L, The effect of 40 Hz light flickering on metabolites involved in SAM-mediated transmethylation, including S-adenosylmethionine (SAM) (I), S-adenosylhomocysteine (SAH) (J), homocysteine (Hcy) (K), and methionine (Met) (L) in V1. *P < 0.05 when comparing 40 Hz with normal light groups at 240 min in (L), **P < 0.01 when comparing 40 Hz group at 30 min or 40 Hz group at 240 min with the groups exposed to normal light (M). M-O, The effect of 40 Hz light flickering on the adenosine degradation product, inosine (by UHPLC-MS/MS analysis; M) and on mRNA expression of adenosine-degrading enzymes adenosine kinase (ADK) (N) and adenosine deaminase (ADA) (O) in V1. *P < 0.05 when comparing the 40 Hz group with the normal light group at 240 min. P-R, The effect of 40 Hz light flickering on the density and mRNA expression of AMPK-α1 (by qPCR analysis) (P-Q) and on AMPK phosphorylation (by Western blot; R) in V1. *P < 0.05 Student’s t-test; comparing the 40 Hz group with the normal light group at 240 min after flickering cessation.

We next explored the contribution of intracellularly generated adenosine to the delayed and sustained rise of extracellular adenosine, which involves an augmented production of intracellular adenosine as a by-product of increased energy (intracellular ATP) consumption followed by an ENT1/2-mediated transmembrane efflux of adenosine. We first assessed the contribution of ENT1/2-mediated transmembrane transport of adenosine after 40 Hz flashing using the ENT inhibitor dipyridamole. We found that 40 Hz light flickering increased the total amount of adenosine (about 90% being intracellular adenosine), as revealed by HPLC analysis (Fig. 4D). Moreover, the increase in the total amount of adenosine induced by 40 Hz flickering was not prevented by pretreatment with dipyridamole (Fig. 4D). Importantly, 40 Hz light flickering induced a robust and sustained increase (up to 2-3 h) in extracellular adenosine levels in V1 of mice treated with vehicle (0.5% methylcellulose), but this increase was almost completely abolished by pretreatment (30 min before light flickering) with dipyridamole (15 mg/kg) (Fig. 4F). Furthermore, when dipyridamole was administered immediately after stopping light flickering delivery, the increase in extracellular adenosine (after an initial increase during the flickering period) was largely prevented, indicating that the sustained increase in extracellular adenosine was also dependent on ENT activity (Fig. 4G). Taken together, the observation that dipyridamole abolished the variation of extracellular adenosine levels but not the total (mainly intracellular) adenosine levels is in line with ENT-mediated adenosine efflux from the intracellular to the extracellular compartments.

The transmembrane transport of adenosine is mainly carried out by two different ENTs, with ENT1 having a relatively higher affinity and lower capacity for adenosine while ENT2 has a relatively lower affinity and higher capacity (*30*). We employed *ENT1*-KO and *ENT2*-KO mice (*26*) to determine their individual contributions to 40 Hz flickering-induced extracellular adenosine generation in V1. As expected, 40 Hz flickering consistently produced a robust increase in extracellular adenosine levels in wild-type (WT) mice. In *ENT1*-KO mice, there was still a significant and sustained increase in extracellular adenosine levels in response to 40 Hz flashing, although slightly lower than in WT mice at later time points (Fig. 4G, insert panel). By contrast, the increase in extracellular adenosine generation during and after light flickering was almost completely abolished in *ENT2*-KO mice (Fig. 4H). These findings underscore the essential role of ENT2, rather than ENT1, in mediating the elevation of extracellular adenosine in the cortex induced by 40 Hz flickering, by promoting the transfer of adenosine from the intracellular to the extracellular compartment.

### 5. 40 Hz flickering induces extracellular adenosine generation resulting from an AMPK-associated control of energy metabolism in V1

We further investigated the specific pathways responsible for intracellular adenosine generation in response to 40 Hz flickering, including stepwise ATP dephosphorylation, SAM-mediated transmethylation, salvage/*de novo* purine synthesis, and impaired adenosine metabolism via adenosine deaminase (ADA) (Fig. 5A). We performed targeted UHPLC-MS/MS and qPCR analyses to specifically assess purine metabolites and the expression of key enzymes involved in purine intracellular metabolism: (a) consistent with the cascade amplification of adenosine generation from stepwise ATP dephosphorylation driven by increased energy consumption, targeted UHPLC-MS/MS analysis revealed a rise in the total concentrations of ADP, AMP, and adenosine without changes in ATP levels in V1 assessed 240 min after 40 Hz flickering stimulation (Fig. 5B to E). There were also no changes in the levels of cAMP and nicotinamide adenine dinucleotide (NAD) (Fig. 5F-G), indicating that these adenosine-generating pathways were not likely to contribute. (b) The targeted UHPLC-MS/MS analysis also revealed a decrease in inosine monophosphate (IMP) levels after 40 Hz flickering stimulation (Fig. 3H), suggesting that the salvage/*de novo* purine synthesis pathway was an unlikely contributor to the increased adenosine generation. (c) Intracellular adenosine can also be generated by the hydrolysis of S-adenosylhomocysteine (SAH) into adenosine and homocysteine via SAH hydrolase (SAHH) within the S-adenosylmethionine (SAM)-dependent transmethylation pathway(*15–17*) (Fig. 5A). The targeted UHPLC-MS/MS analysis revealed no changes in SAM, SAH, or methionine associated with increased homocysteine levels after 40 Hz flickering (Fig. 3I to L). Thus, it is unlikely that the increased generation of intracellular adenosine is due to the involvement of S-adenosylmethionine (SAM)-dependent transmethylation. (d) The UHPLC-MS/MS analysis showed elevated levels of inosine (Fig. 5M) and qPCR analysis revealed an increase in adenosine deaminase (ADA) mRNA, but not astrocytic adenosine kinase (ADK) mRNA (Fig. 5N to O), suggesting a compensatory increase in adenosine metabolism through deamination of adenosine to inosine via ADA (Fig. 3R) and not through phosphorylation of adenosine to AMP through ADK (Fig. 3Q). This indicates that reduced adenosine metabolism is not a contributing factor to intracellular adenosine accumulation after 40 Hz flickering. Taken together, the UHPLC-MS/MS and qPCR analyses suggest that the increased intracellular adenosine generation induced by 40 Hz flickering is mainly associated with the stepwise ATP dephosphorylation for energy metabolism, rather than SAM-mediated transmethylation or S-adenosylhomocysteine hydrolysis, or the salvage/*de novo* purine synthesis pathways, or the impaired adenosine metabolism via ADA. Altered energy metabolism with increased production of endogenous adenosine is critically dependent on AMPK, a key metabolic controller (*31*). To further confirm the involvement of increased energy metabolism in extracellular adenosine generation, we investigated the frequency (20 Hz, 40 Hz, and 80 Hz) and time (30 and 240 min after exposure) dependence of light flickering on phosphorylated AMPK in V1 and AMPK mRNA by qPCR analysis. After 40 Hz light flickering, we observed an increase in AMPK-α1 mRNA, but not AMPK-α2, in V1 at 240 min compared to control mice exposed to normal light (Fig. 5P to Q). Similarly, 240 min after light flickering cessation, we observed a significant increase in phosphorylated AMPK in response to 40 Hz light flickering (Fig. 5R). Thus, the increased expression and phosphorylation of AMPK may contribute to the delayed and sustained increase in extracellular adenosine levels after light flickering.

### 6. Brief 40 Hz light flickering promotes sleep onset and maintenance without affecting slow-wave activity power in mice

To consolidate the adenosine hypothesis as the neurochemical basis for the biological effects of 40 Hz light flickering, we evaluated the effect of 40 Hz visual pulses on sleep, since adenosine is a well-known physiological regulator of homeostatic sleep needs. On day 1, mice were exposed to normal light, and we recorded EEG/EMG/locomotion in mice throughout the dark phase (7:00 pm-7:00 am). On day 2, during the 7:00 am-7:00 pm light cycle, we exposed mice to 40 Hz light flickering in the last 30 min of the light phase (6:30-7:00 pm) and then recorded EEG/EMG/locomotion during the subsequent dark phase. By comparing the sleep duration on day 2 to that of day 1, we observed a significant increase in the sleep amount induced by 40 Hz flickering. This sleep-promoting effect was prominent in the initial 2-3 h of the dark phase but not for the remainder period. During the first 3 h of the dark phase, the previous 40 Hz flickering significantly increased the total duration of sleep by enhancing both slow-wave sleep (SWS) (increased by 2.2-fold, mean ± SEM, from 19.92 ±3.4 min to 63.97 ±3.67 min) and rapid-eye-movement sleep (REMS) (increased by 3.2-fold, mean ±SEM, from 1.13 ±0.66 min to 4.83 ±0.90 min), compared to mice exposed to normal light (Fig. 6A to E). Moreover, we confirmed the frequency specificity of the somnogenic effect of light flickering by demonstrating that exposure to visual flashing at 20 Hz or 80 Hz did not alter the sleep pattern (Fig. 6F to H). The detailed analysis of sleep amount at 30-min intervals revealed that the effect of light flickering frequency on sleep was not immediate but became evident and sustained for 2 h after flickering cessation (Fig. 6G). These findings are consistent with the frequency-specific effect on extracellular adenosine generation (Fig. 1B). The correspondence between the frequency-specific effect on extracellular adenosine production and sleep amount suggests that the sleep-promoting effect of 40 Hz flickering is due to increased extracellular adenosine signaling in the brain. Notably, there was no significant difference in the delta power density of SWS during the dark phase between mice exposed to normal light and those exposed to 40 Hz flickering (Fig. 6I). This observed lack of an impact of 40 Hz light flickering on SWS delta power density suggests that it may be a potential treatment for insomnia, as it promotes sleep without side effects of daytime fatigue and drowsiness.

**Fig. 6.**
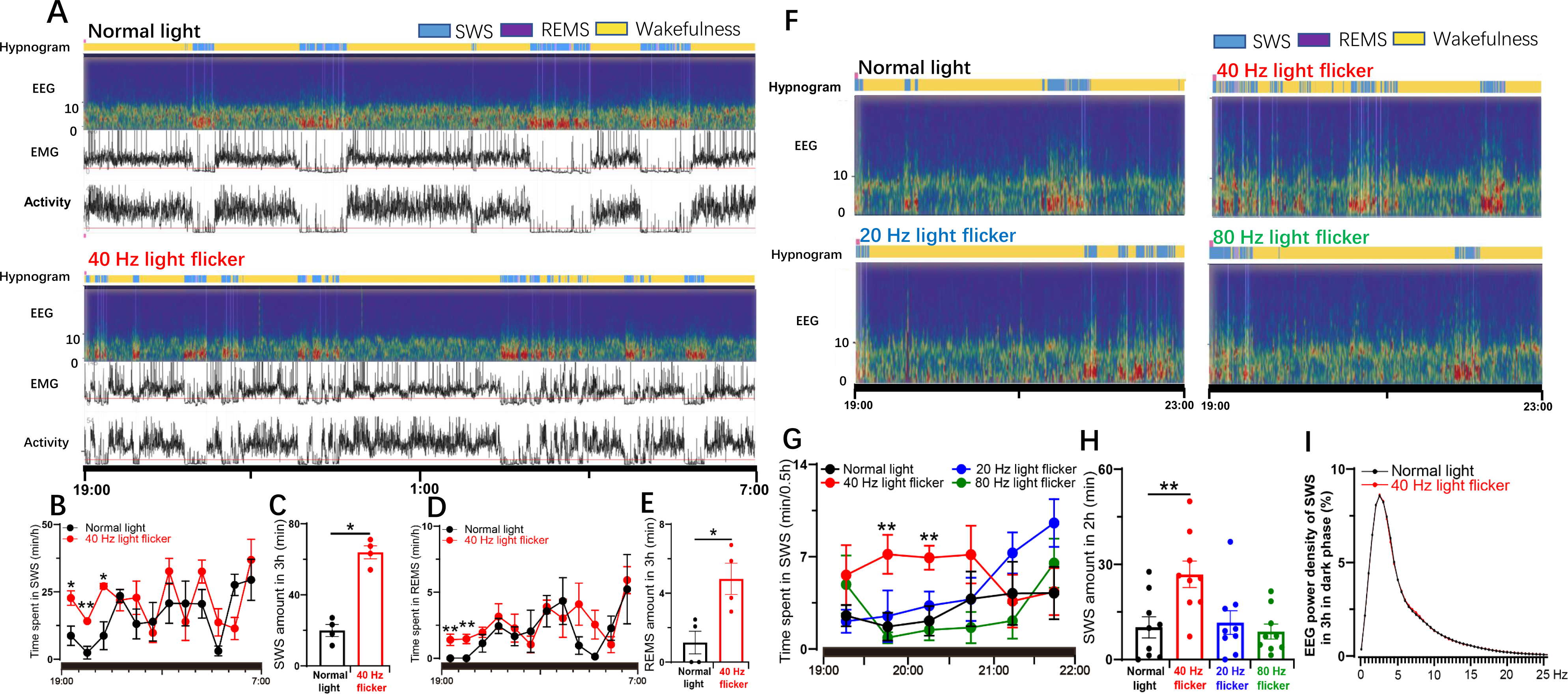
Light flickering at 40 Hz induces sleep in a frequency-dependent manner. A, The hypnogram, EEG spectrum, EMG and locomotor activity in mice exposed to normal light and 40 Hz flickering, respectively. B and D, The time-course of SWS and REMS in mice throughout the dark phase after exposure to normal light or 40 Hz light flickering treatments 30 min before the dark phase, respectively (n = 4/group); *P < 0.05; **P < 0.01, mean ± SEM, 40 Hz vs. normal light, assessed by two-way ANOVA. C and E, The amount of SWS and REMS during the initial 3 h after exposure of mice to normal light or 40 Hz light flickering 30 min before the dark phase (n = 4/group); *P < 0.05, 40 Hz vs. normal light, assessed by Student’s t-test. F, Representative hypnograms and EEG spectra of mice after treatments with light flickering at 20 Hz, 40 Hz, and 80 Hz, respectively. G, The time-course of SWS during 30 min blocks in the first 3 h after illumination at 20 Hz, 40 Hz, and 80 Hz frequencies. **P < 0.01 indicating significance at 20 Hz, 40 Hz, and 80 Hz compared to normal light using Student’s t-test (n = 9/group). H, The amount of SWS and REMS during the following 2 h after exposure to normal light or 20, 40, and 80 Hz light flickering treatments 30 min before the dark phase (n = 9/group). I, Superimposable power density of the EEG spectra in mice after either normal light or 40 Hz light flickering treatments (n = 4/group).

### 7. The 40 Hz flickering-induced sleep-promoting effect requires ENT2 but not ENT1 activity

To further confirm the crucial role of extracellular adenosine signaling via ENT2 in mediating the sleep-promoting effect of 40 Hz flickering, we conducted sleep profile recordings (EEG and EMG) in mice deficient in either ENT1 or ENT2 after exposure to 40 Hz flickering for 30 min. In WT mice (*ENT2*-KO littermates), 40 Hz flickering stimulation increased SWS amount during the first 2 h by 1.1-fold (from 11.77 ±2.59 min to 24.65 ±4.49 min, mean ±SEM) (Fig. 7A and B). However, the sleep-inducing effect of 40 Hz light flashing was abolished in *ENT2*-KO mice and even lower than the normal light treatment, indicating that ENT2 is critical not only for mediating the sleep-inducing effect of 40 Hz light flickering but also for spontaneous sleep/wakefulness regulation (Fig. 7C and D). In contrast, 40 Hz flickering still significantly increased SWS in *ENT1*-KO mice, with the total amount of sleep increasing by 1.5-fold (from 25.04 ±8.01 min to 62.86 ±13.26 min, mean ± SEM) during the first 4 h (Fig. 7E and F), consistent with the lack of effect of *ENT1*-KO on extracellular adenosine levels. These findings support the conclusion that extracellular adenosine generation via ENT2, but not ENT1, activity is required for the sleep-promoting effect of 40 Hz light flickering.

**Fig. 7.**
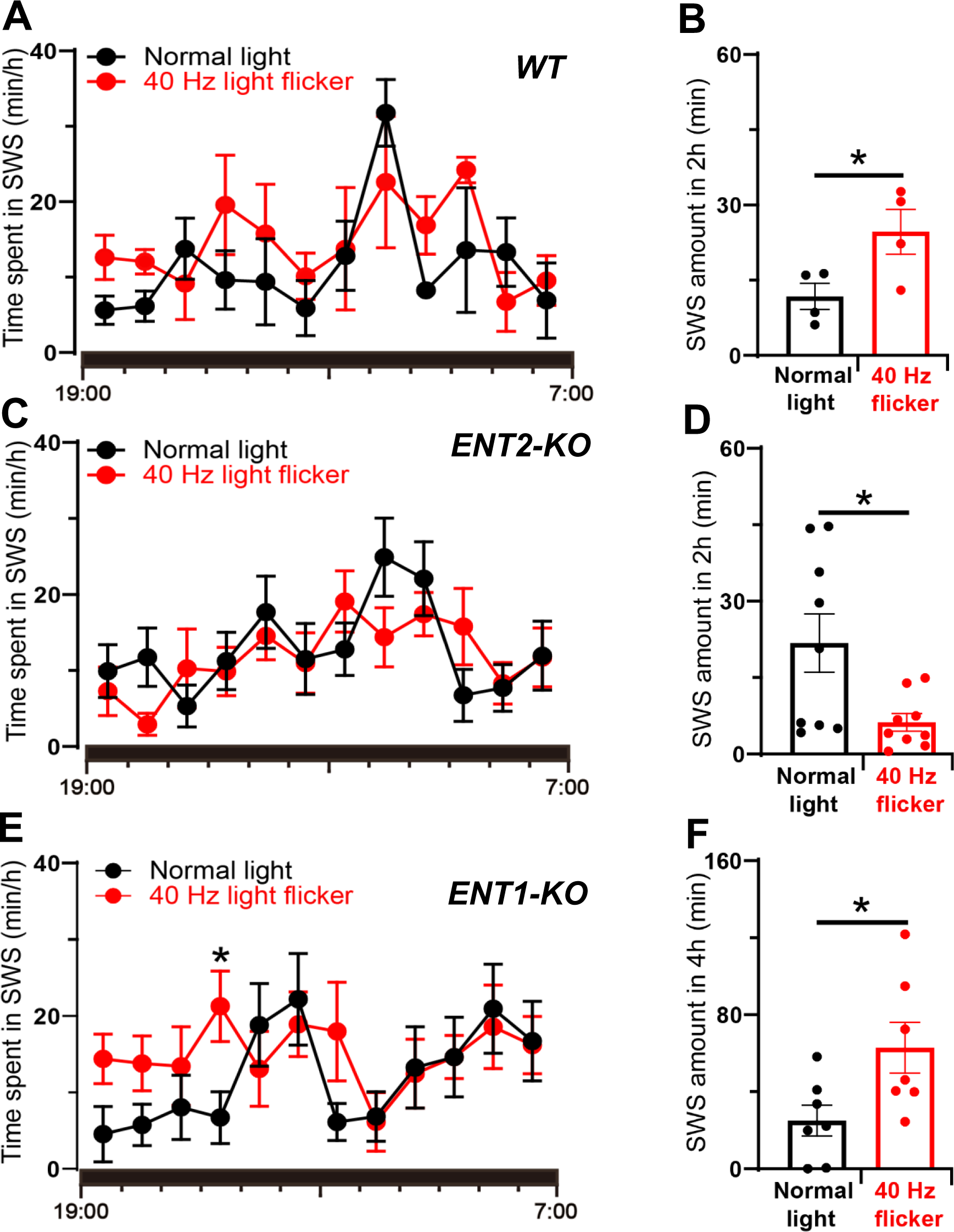
The sleep-promoting effect of 40 Hz light flickering is mediated by ENT2, but not ENT1, in mice. A-B, WT littermates were exposed to normal light (day 1) and 40 Hz light flickering (day 2) for 30 min before the dark phase. The time-course of slow wave sleep (SWS)during the entire dark phase after light flickering was recorded and analyzed in WT littermates. The amount of SWS in the first 2 h was compared between mice subjected to 40 Hz flickering and normal light, and statistical significance (*P <0.05) was evaluated using Student’s t-test (n = 4). C-D, *ENT2*-KO mice were exposed to normal light (day 1) and 40 Hz light flickering (day 2) for 30 min before the dark phase. The time-course of SWS during the entire dark phase after light flashing was recorded and analyzed in *ENT2*-KO mice. The amount of SWS during the first 2 h in *ENT2*-KO mice was compared between 40 Hz flickering and normal light conditions by Student’s t-test (*P <0.05; n = 10/*ENT2*-KO). E-F, *ENT1*-KO mice were exposed to normal light (day 1) and 40 Hz light flickering (day 2) for 30 min before the dark phase. The time-course of SWS during the whole dark phase after light flickering was recorded and analyzed in ENT1-KO mice. The amount of SWS in the first 4 h in ENT1-KO mice was measured, and the statistical significance of the difference between 40 Hz flickering and normal light was evaluated by Student’s t-test (*P <0.05; n = 7/ENT1-KO). The time-course of SWS x KO interactions was analyzed using two-way ANOVA, while the difference in the total sleep amount each hour was analyzed with a paired t-test. *P < 0.05 was considered significant. Data are presented as mean ± SEM.

### 8. Focal injection of adenosine into the primary visual cortex increases SWS dose-dependently

To investigate the neuronal circuit/locus responsible for the sleep-promoting effect of 40 Hz light flickering, we probed whether local adenosine administration into the visual cortex could replicate the sleep-promoting effect of 40 Hz flickering. Following the implantation of the injection cannulas for ten days, we microinjected adenosine or artificial cerebrospinal fluid (ACSF) into V1 on days 11, 14, and 17 at 6:30 pm. Either ACSF or adenosine at a dose of 1.5 or 4.5 nmol/side was injected into V1 immediately before the dark phase onset. We then monitored the sleep patterns of the mice for the next 12 h, from 7:00 pm to 7:00 am. The analysis showed that the injection of adenosine at both doses resulted in a significant increase in the amount of SWS (ACSF group: 52.11 ±5.11 min; adenosine 4.5 nmol/side: 96.24 ±6.14 min; adenosine 1.5 nmol/side: 67.64 ± 3.18 min; mean ± SEM) during the first 3 hours, compared to the mice injected with ACSF (Fig. 8A to C). Furthermore, the focal adenosine-induced sleep effect was dose-dependent, with the greatest effect observed at the higher dose of 4.5 nmol/side, and a moderate effect observed for the lower dose of 1.5 nmol/side (Fig. 8A to C). These data provide direct evidence that the local elevation of extracellular adenosine levels in V1 is sufficient to promote SWS and that the visual cortex plays a crucial role in mediating 40 Hz flickering-induced sleep via adenosine signaling.

**Fig. 8.**
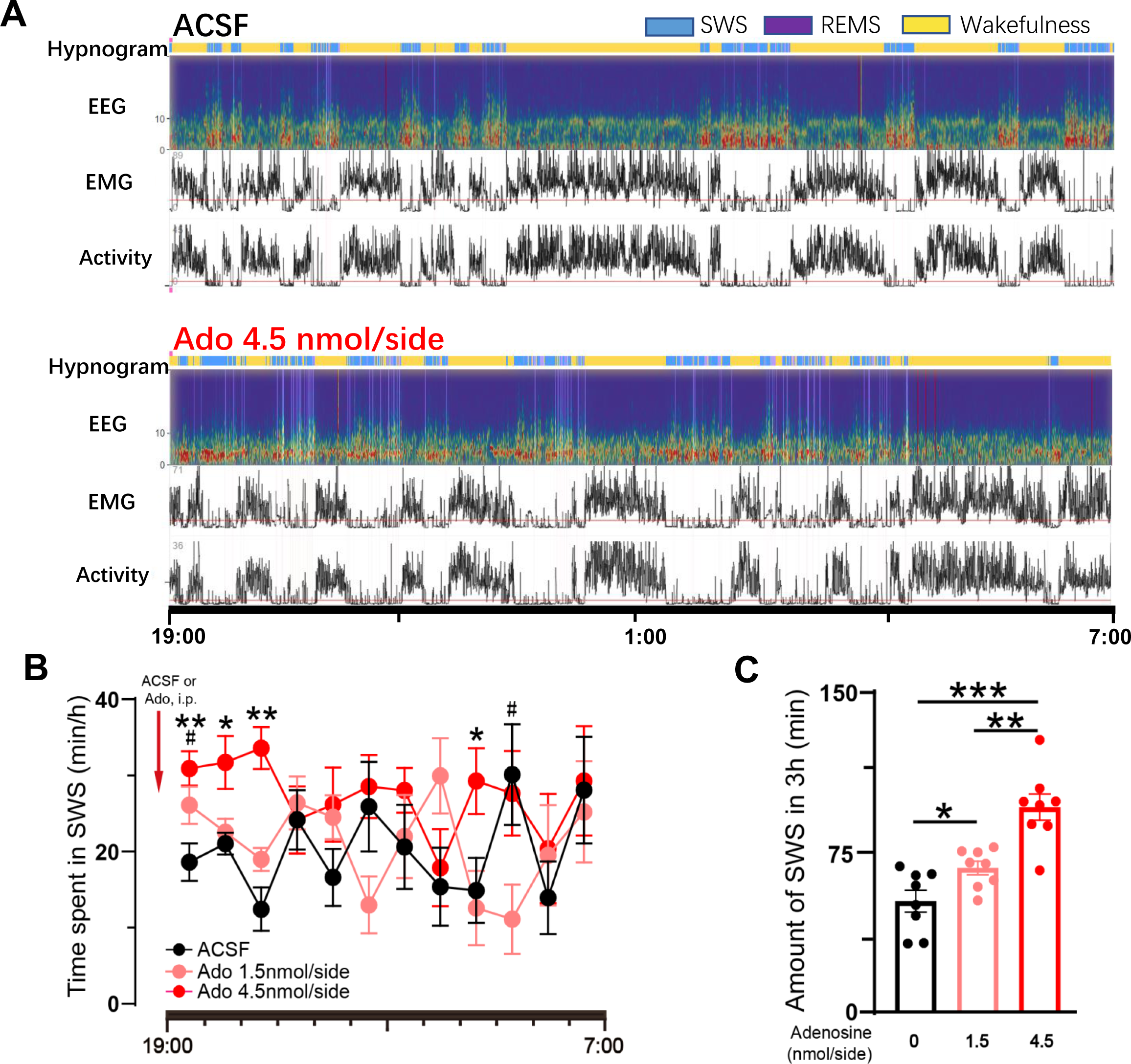
Bilateral microinjection of adenosine into the visual cortex increases slow-wave sleep in a dose-dependent manner. A, Representative hypnograms, EEG spectra, EMG and locomotor activity recordings in mice after the administration of vehicle (ACSF) and adenosine (Ado, 4.5 nmol/side) in the visual cortex. B, The time-course of SWS in WT mice following bilateral microinjections of ACSF or adenosine at doses of 1.5 and 4.5 nmol/side into V1. Statistical significance was assessed by two-way ANOVA for the time course (time x adenosine interaction effect), followed by LSD post-hoc comparison. *P < 0.05 and **P < 0.01 for the 4.5 nmol/side adenosine group compared with the ACSF group, and #P < 0.05 for the 1.5 nmol/side adenosine group compared with the ACSF group. C, The amount of SWS during the first 3 h after bilateral microinjections of ACSF or adenosine at doses of 1.5 and 4.5 nmol/side into V1. Statistical significance was evaluated by one-way ANOVA followed by the LSD test (n = 8/group); *P < 0.05 and **P < 0.01 were considered significant.

### 9. 40 Hz light flickering reduces sleep latency and wake time after sleep onset while increasing total sleep time and efficiency in children with insomnia

Finally, we conducted a self-controlled clinical study to assess the effectiveness of 40 Hz light flickering in promoting sleep onset and maintenance in 49 children with insomnia symptoms, with an average age of 9.63 ± 3.28 years and body-mass index (BMI) of 15.60 (14.70-18.00) kg/m^2^, both sexes balanced (44.90 % female) (Figure 9A and B). The patients underwent sleep adjustment at the sleep center and their sleep parameters (including EEG and EMG) were monitored over three days. On the first day, sleep parameters were assessed for the patient’s baseline activities. On the second day of the study, the patients were exposed to 40 Hz light flickering for 30 min (8:30-9:00 pm) and their sleep parameters were recorded during the entire night. Analysis of the primary endpoint by comparing day 2 (40 Hz light flickering) with day 1 (baseline with normal light exposure) using polysomnography analysis revealed that 40 Hz light flickering significantly reduced sleep onset latency (SOL), which is the duration from light off to sleep onset (W = -767, P < 0.001), as depicted in Fig. 9C and Table 1. Furthermore, analysis of the secondary endpoints using polysomnography (PSG) also showed that 40 Hz light flickering had a positive effect on total sleep time (TST) (W = 905, P < 0.001, Fig. 9D and Table 1). Sleep efficiency (SE) was similarly enhanced by 40 Hz light flickering (W = 1055, P < 0.001, Fig. 9E and Table 1). Light flickering at 40 Hz reduced wake time after sleep onset (WASO, minutes of wake after sleep onset) (W = -924, P < 0.001, Fig. 9F and Table 1). We noted that 40 Hz light flickering had no significant effect on arousal frequency (AF, defined as the number of awakenings during the night; t(_48_) = 1.649, P = 0.106; Fig. 9G and Table 1). In addition, 40 Hz light flickering also shortened REM sleep onset latency (REM SOL, measured in minutes between sleep onset and the first epoch of REM sleep) (t(_48_) = 4.521, P < 0.001, Fig. 9H and Table 1). Notably, the percentage of light sleep (Fig. 9I and Table 1) or deep sleep (Fig. 9J and Table 1) remained unchanged after 40 Hz light flickering (W= -322, P = 0.100; t(_48_) = 0.510, P = 0.612). We found that 40 Hz flickering increased REM sleep percentage (W = 447, P = 0.025, Fig. 9K and Table 1). Taken together, in this clinical study involving 49 children with insomnia symptoms, 40 Hz light flickering for 30 min (between 8:30 pm and 9:00 pm) was found to enhance sleep onset and maintenance. This was evidenced by a reduction in the time required to fall asleep, an increase in total sleep time and sleep efficiency, and a reduced wake time after sleep onset.

**Fig. 9.**
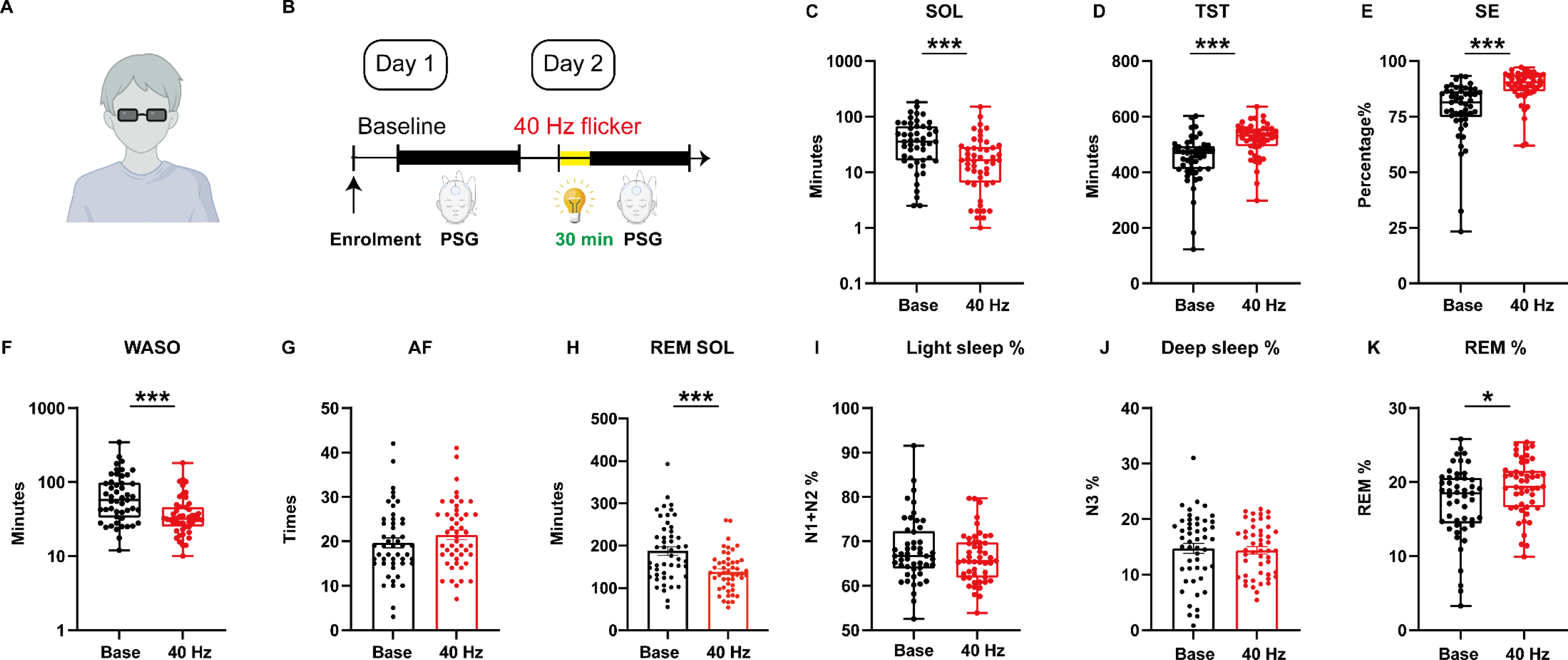
40 Hz light flickering improves sleep onset and maintenance among children with insomnia. (A) Schematic figure for 40 Hz flickering stimulation. (B) Illustrations of the study design, in which patients were enrolled for three consecutive days: day 1 served as a baseline, day 2 involved 30 min of 40 Hz flickering stimulation before sleep. (C) 40 Hz flickering significantly reduced SOL (W = -767, P < 0.001). D. 40 Hz flickering significantly increased TST (W = 905, P < 0.001). E. Sleep efficiency (SE) was also improved by 40 Hz flickering (W = 1055, P < 0.001). F. WASO was remarkably reduced by 40 Hz flickering stimulation (W = -924, P < 0.001). G. Arousal frequency (AF) remained unchanged (t(48) = 1.649, P = 0.106) after 40 Hz flickering stimulation. H. REM SOL was also reduced by 40 Hz flickering (t(48) = 4.521, P < 0.001). The percentage of light (I) and deep sleep (J) remained unchanged (W= -322, P = 0.100; t(48) = 0.510, P = 0.612). An increase in REM sleep percentage (K) was detected (W = 447, P = 0.025) after 40 Hz flickering stimulation. Paired t test or Wilcoxon matched-pairs signed rank test was conducted based on the distribution types of the paired difference. PSG, Polysomnography; SOL, sleep onset latency; TST, total sleep time; SE, sleep efficiency; WASO, wake time after sleep onset; AF, arousal frequency; REM SOL, REM sleep onset latency; REM, rapid eye movement; N1 + N2, light sleep; N3, deep sleep. *P < 0.05, **P < 0.01, ***P < 0.001, ns: not significant; data are presented as mean ± SEM or median (P25-P75).

**Table 1.**
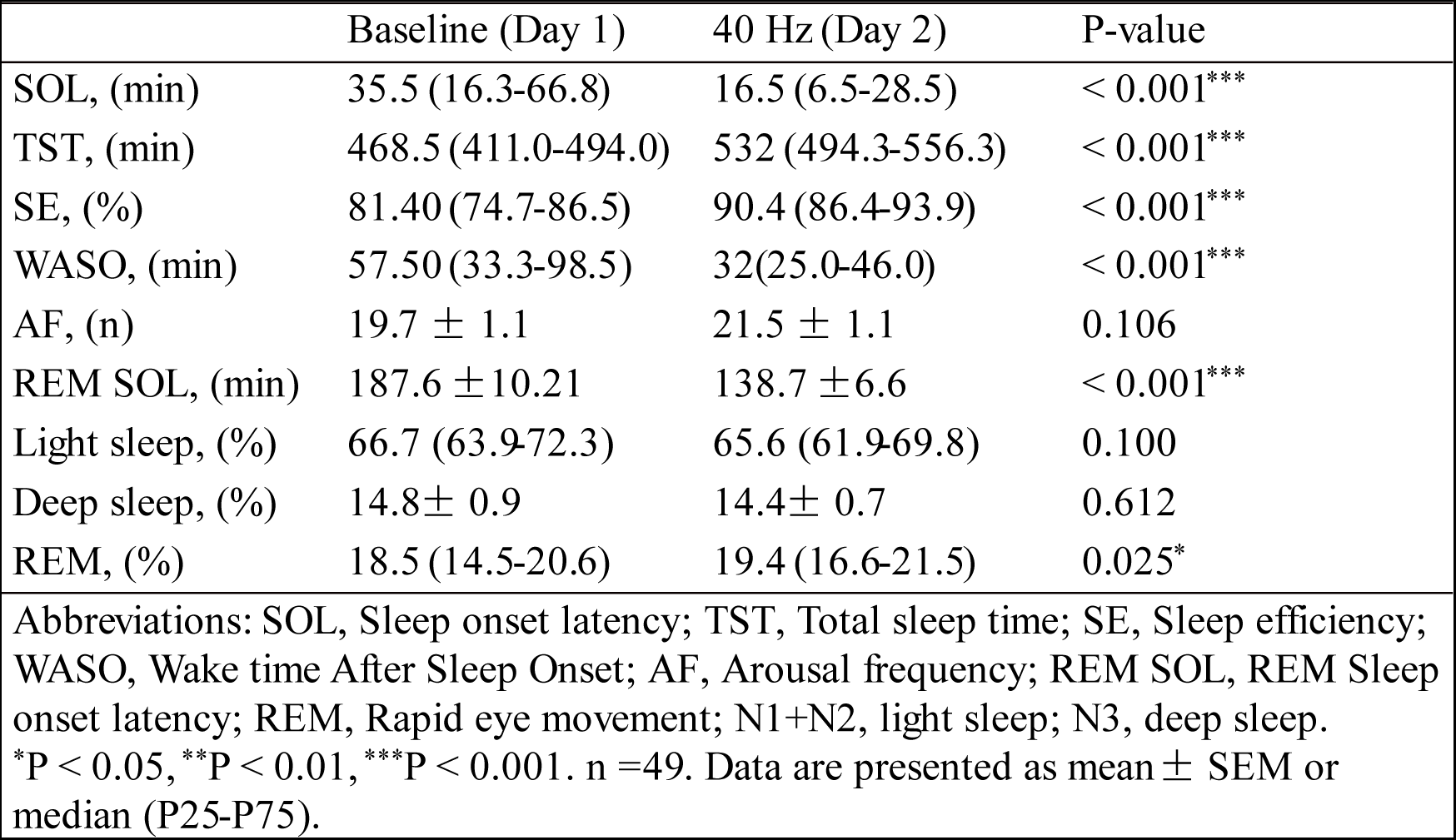
Effects of 40 Hzflickeronsleep parameters in children with insomnia symptoms.

## Discussion

Our findings establish cortical adenosine signaling as the neurochemical basis responsible for the sleep-inducing effects of 40 Hz flickering, reinforcing its therapeutic potential for brain disorders. The neurochemical underpinning of this phenomenon is primarily attributed to the intrinsic photo-electro-chemical coupling between light flickering-triggered neural activities (such as gamma oscillation) and the energy-demanding metabolic processes that lead to heightened energy metabolism and the subsequent adenosine generation as dictated by the [ATP]/[ADP][AMP] ratio (*15*). Accordingly, we discovered that 40 Hz light flickering resulted in robust LFP responses and a significant, sustained increase in extracellular adenosine levels in the primary visual cortex. This intrinsic relationship aligns with previous findings that specific visual pattern stimuli selectively enhance LFP oscillations in the gamma range, which is tightly linked to and limited by mitochondrial activity in the brain (*12–14*) and increased regional cerebral blood flow in healthy individuals (*7, 32, 33*) and (*12–14*). Notably, the observed sustained increase in AMPK phosphorylation (a key sensor and controller of cellular energy metabolism formatting sleep quality (*34, 35*)), which lasts up to 4 hours, is of particular interest as it may underlie the persistent increase in cellular energy metabolism and of the extracellular levels of adenosine (*36, 37*) after light flickering cessation. This neurochemical underpinning involves the role of extracellular adenosine as a “retaliatory metabolite”, which serves as a connection between neurotransmission, cellular metabolism, and tissue responses to light (*15, 16*). The sustained increase in extracellular adenosine levels not only plays a neuromodulatory role in regulating synaptic information flow (*18, 19*) but also appears to be crucial for maintaining cellular homeostasis by controlling the availability and utilization of oxidative substrates in intermediary metabolism and providing protective effects (*18, 19*). These neurochemical and molecular adaptations beyond gamma oscillations observed after 40 Hz visual flickering, including the sustained increase of extracellular adenosine for 3 h after flickering cessation, play a critical role in producing long-lasting therapeutic effects on pathological changes. Collectively, the intrinsic link between the high cellular metabolism imposed by gamma oscillation and adenosine generation, together with the multiple physiological effects of adenosine as a dual neuromodulator, homeostasis regulator, uniquely position the adenosine signaling system to serve as the unique and pivotal neurochemical basis for the biological effects of 40 Hz light flickering. This identification of extracellular adenosine as the molecular and neurochemial underpinning of 40 Hz flickering fills the critical gap between bolstered neuronal activity (specifically gamma oscillation) and therapeutic effects such as beta-amyloid reduction and cognitive improvements that are controlled by the adenosine modulation system (*18*).

The present discovery that 40 Hz light flickering can specifically evoke an endogenous homeostatic regulatory mechanism operated by increased adenosine signaling, provides a novel, promising and non-invasive approach to insomnia treatment. In fact, adenosine is an endogenous somnogen factor driving both homeostatic sleep needs (*20–22, 24*) and potentially contributing to motivation-related arousal (*20, 38, 39*). However, the development of adenosine-based therapies for insomnia has been largely limited by the adverse cardiovascular and liver effects associated with the widespread distribution of adenosine receptors and the challenges in transporting across the blood-brain barrier to most drugs targetting the adenosine modulation system (*16, 19*). Through its neurochemical coupling mechanism, 40 Hz light flickering enhances adenosine signaling in the brain, conferring a novel and non-pharmacological strategy to treat insomnia. Unlike traditional anti-insomnia drugs, the treatment of isomnia by 40 Hz light flickering is characterized by being non-invasive and rapidly effective, requiring only 30 min before sleep, by its potent somnogenic effect, leading to a 2.2-fold increase in SWS duration in mice. This light therapy promotes both the onset and maintenance of sleep, as indicated by decreased wake-after-sleep-onset and increased total sleep time, along with its favorable risk-to-benefit ratio and low cost. Furthermore, this treatment improves sleep amount without affecting delta power density in mice or the ratio of deep to light sleep in children with insomnia, leading to improved sleep efficiency and without significant sleep rebound or side effects (*20, 40*). Notably, 40 Hz light flickering promotes sleep by activating adenosine signaling in the visual cortex, as the injection of adenosine into the visual cortex reproduces the somnogenic effects of 40 Hz light flickering. The critical role of adenosine signaling in the visual cortex in promoting sleep is further supported by recent studies revealing shared and structured global patterns of cortical activity (*41, 42*). Moreover, optogenetic inhibition of the REM-like pattern in the occipital cortex can enhance SWS and block the transition from SWS to REM sleep in mice (*41, 42*). Thus, adenosine signaling serves as a gateway for both physiological and pharmacological transitions from wakefulness to sleep by directly modulating activity in the visual cortex. The present study also identified ENT2 as a key molecular regulator of the therapeutic effects of 40 Hz light flickering effect on sleep control since the genetic deletion of ENT2, but not ENT1, abolished the frequency-specific effects of light flickering on both extracellular adenosine generation and the induction and maintenance of sleep. The critical role of ENT2 in the generation of extracellular adenosine matches recent findings that neuronal activity-induced outflow of adenosine from neurons after high-frequency stimulation in the hippocampus is ENT- dependent (*26, 27, 43*). Notably, the greater importance of ENT2 compared to ENT1 may be attributed to its larger capacity to transport adenosine, with four times higher expression in neurons and astrocytes, allowing larger increases of extracellular adenosine levels in conditions such as light flickering (*30, 44*).

The identification of adenosine signaling as a neurochemical coupling mechanism for 40 Hz flickering also provides a rationale to explain its therapeutic effects in AD and ischemic pathologies. As an illustration, adenosine signaling can activate microglial cells (*5, 6*) and enhance vascularization in various tissues (*7*), including our recent demonstration that the A_2A_ receptor controls microglial activation (*18, 45*) and retinal vascularization (*46, 47*). The induction of adenosine signaling through 40 Hz flickering is expected to boost microglial activation and vascularization in the cortex, increasing the clearance of beta-amyloid species and improving cognition in AD mice. Furthermore, the protective effect of light flickering at 30-50 Hz against ischemia-induced hippocampal neuronal death and disruption of hippocampal CA1 low gamma oscillations (*8*) can be partially attributed to increased extracellular adenosine levels, which suppress excessive excitatory transmission by acting as a gatekeeper mechanism via the adenosine A_1_ receptor (*18*). Thus, this study reveals a novel and non-invasive treatment for insomnia with potentially wider therapeutic implications for other neuropsychiatric disorders, attributed to the dual roles of adenosine as both a neuromodulator and homeostatic regulator.

## Materials and methods

### Animals

All experimental protocols (#wydw2021-0563) were approved by the Institutional Ethics Committee and followed the guidelines for Animal Use in Research and Education at Wenzhou Medical University, China. C57BL/6 wild-type mice were purchased from institute-approved vendors (Beijing Vital River Laboratory Animal Technology Co., Ltd). We obtained vGluT2- IRES-CRE mice (Jackson stock #: 016963) and CD73 knockout (*CD73*-KO) mice (Bar Harbor, ME) from Jackson Laboratory. *ENT1*-KO mice, *ENT2*-KO mice, and GRAB_ATP1.0_ knock-in (GRAB_ATP1.0_-KI) mice were generated using CRISPR/Cas9 technology by Beijing Biocytogen and kindly provided by Dr. Yulong Li’s laboratory. The mice were housed in an enriched environment with *ad libitum* access to food and water under a 12 h light/12 h dark cycle. Mice with implants for EEG/EMG recording, optogenetic manipulation and fiber photometry recording were housed individually.

### Surgery: viral injection and cannula implantation

The mice were anesthetized with isoflurane and head-fixed in a stereotaxic apparatus (Kopf, USA) to expose the skull and drill a hole for injection and implantation. (i) A craniotomy was performed on top of V1 and a 0.5-1 mm-thick glass pipette was used to inject (20 nL/s) virus (0.2 - 0.4 µL) into V1 at coordinates AP: -3.6 mm, ML: ±2.4 mm, DV -0.6 mm from the cortical surface using KDS LEGATO 130 (KD Scientific, China). The following AAV viruses were used in the current study: AAV2/9-hSyn-GRAB_Ado1.0_, AAV2/9-GfaABC1D-GRAB_Ado1.0_, AAV2/9-hsyn-GRAB_Ado- mut_, AAV2/9-hSyn-DIO-ChrimsonR-mCherry, AAV2/9-hSyn-ChrimsonR-tdTomato, AAV2/9- EF1a-flex-taCasp3-TEVp, AAV2/9-hSyn-taCasp3-T2A-TEVp, AAV2/9-hSyn-CRE, AAV2/9- mDlX-CRE, AAV2/9-hSyn-tdTomato, AAV2/9-hSyn-DIO-mCherry, and AAV2/9- GfaABC1D-mCherry-hPMCA2wb. (ii) After viral injection, a 0.5-1 mm diameter craniotomy was performed to implant one or two optical fibers with FC ferrules (200 µm diameter, Thorlabs, USA) into V1 at the same coordinates as the virus injection for conducting fiber photometry recordings or optogenetic manipulation experiments. (iii) For EEG recording, four stainless steel screws were implanted, with two located over the primary motor cortex (approximately 2 mm anterior to the bregma and 1 mm lateral to the midline) and the other two over the parietal cortex (approximately 2 mm posterior to the bregma and 1 mm lateral to the midline). (iv) For EMG recording, two stainless steel wires were implanted in the neck muscle. After attaching both EEG and EMG electrodes to a head-mounted device (Catalog #2631, Bio-Signal Technologies, China), the assembly was secured with dental cement. (v) To inject adenosine through the implanted cannula, two 26-gauge stainless steel guide cannulas (62003, RWD Life Science, China) were stereotaxically implanted into V1 at AP -3.6 mm, left-right ±2.4 mm from the bregma, and DV - 0.7 mm from the skull top. Matched dummy cannulas (62003, RWD Life Science, China) were inserted into the guide cannulas after surgery and removed only during microinjections. Microinjections of ACSF or adenosine solutions were performed using a tubing-nested 10-µL Hamilton syringe (80365, Hamilton, USA). After all surgical procedures, mice were housed individually in clear acrylic cages under controlled temperature, humidity, and lighting conditions (12-h light/12-h dark schedule), with *ad libitum* access to food and water.

### *In vivo* electrophysiology

#### Surgery

C57/BL6 mice aged 8-10 weeks were implanted with electrodes to record LFPs in V1. The surgical procedure, as previously described (*48*), involved anesthesia with isoflurane, head fixation in a stereotaxic apparatus (Kopf, USA), and exposing the skull. A cranial window (V1: AP: -3.6 mm, ML: ±2.4 mm, DV: -0.6 mm) was then prepared using a dental drill (RWD Life Science, China) on the left side for implantation. A 16-channel microwire array electrode (Catalog #KD-MWA-16, Kedou, China) was inserted into V1 and grounded above the cerebellar hemispheres using two bone screws (Catalog #700-00122-00, RWD, China). The electrode was then secured in place with tissue glue (Catalog #454, LOCTITE, Ireland) and dental cement (Catalog # type II 2#, New Century Dental, China).

#### Recording

After a recovery period of 14 days following surgery, the mice were transferred to an electrophysiological recording chamber and allowed to acclimate for 20 min before recording began. When the experiment started, the mice were placed in a transparent and electromagnetically shielded box. Each mouse was connected to the recording system via Cereplex M (Cerebus, Blackrock Microsystems, USA), a 16-channel digitally programmable amplifier filtered (0.3 Hz - 7.5 kHz), close to the animal’s head. The signals were sampled at 1000 Hz and refined with a low-pass filter at 250 Hz. The lamps generating different visual stimulation (DC, 10 Hz, 20 Hz, 40 Hz, and 80 Hz) were positioned face to face in front of the box. We isolated the stimulation and recording circuits to avoid any electromagnetic interference. We confirmed that there were no changes in the power of LFPs during visual stimulation in recordings from saline, dead mice (electrode implanted in V1), and negative contrast (electrode implanted in the primary motor cortex (M1)), which helped us exclude any potential photoelectric effect or other interference (data not shown). LFPs were initially recorded in V1 for 5 min under quiet and dark conditions, followed by 30 min of recording during visual stimulation with different frequencies (presented in random order for each mouse), and finally stopped after a 5-min post-stimulation period.

#### Analysis

The LFP data underwent a short-time Fourier transform with MATLAB R2020a and the average gamma power (22-90 Hz)(*13*) or power within flickering frequency (specific frequency ± 0.5Hz) was calculated by integrating the power spectral density (PSD) estimate using the rectangle method. The mean gamma power of DC was used for normalization.

### Light flickering stimulation

Light flickering stimulation was performed using a previously described method (*5–7*). Before the experiment began, the mice were placed in a PVC case, similar to their home cage but without bedding, and kept under dim lighting for 1 h. Two LED bulbs were positioned on the long side of the cage opposite each other and controlled by a single circuit-control relay in a dark room. White LEDs, which emit a combination of visible wavelengths ranging from 390 to 700 nm, were used to provide five types of stimulation (dark, light, 20 Hz, 40 Hz, and 80 Hz) for 30 min, with stimulation consisting of 12.5 ms light on and 12.5 ms light off and using 60 W. The LEDs used in the experiment had a correlated color temperature (CCT) of 4000 K and were operated at various intensities, ranging from 50 to 3000 lux, including 50 lux, 500 lux, 1000 lux, 2000 lux, and 3000 lux. To assess the effect of different duty cycles on extracellular adenosine levels, we conducted explicit tests using a 40 Hz and 3000 lux stimuli with duty cycles of 1%, 5%, and 10%.

Moreover, we examined the effects of light flickering at different wavelengths (380 nm, 460 nm, 520 nm, 650 nm, and 850 nm) compared to a white light flicker at 40 Hz and 500 lux stimulus to investigate their influence on increasing extracellular adenosine levels in V1.

### Preparation and administration of drugs

Dipyridamole was dissolved in a 0.5% methylcellulose solution at 1.5 mg/mL. Felodipine was dissolved in 2% DMSO with normal saline (0.9% NaCl) and covered to avoid light exposure. Nimodipine (felodipine) and the vehicle (2% DMSO in saline) were intraperitoneally injected (i.p.) at a dose of 10 mg/kg body weight before 40 Hz flickering exposure. To prepare the cannula microinjection solution, adenosine was dissolved in ACSF (containing in mM: 126 NaCl, 26 NaHCO_3_, 10 glucose, 2.5 KCl, 1.25 NaH_2_PO_4_, 2 MgCl_2_, and 2 CaCl_2_; pH 7.4) at the required doses (1.5 and 4.5 nmol/side) with an injection volume of 2 µL per side. All drug solutions were prepared approximately 30 min before injection. The injections were performed using a crossover design.

### Fiber photometry recording and analysis

#### Fiber photometry recording during flickering visual stimulation

To record fluorescence from the adenosine sensor or ATP sensor, we attached an optical fiber (Thorlabs, FT200UMT, USA) to the implanted ferrule through a ceramic sleeve and recorded the emission fluorescence using a commercial fiber photometry system (Thinker Tech Nanjing Biotech CO., Ltd). Fiber recordings were performed on freely moving mice during flicker visual stimulation, and no data were excluded. The signal from each continuous experimental trial was normalized to the average fluorescence using a MATLAB program developed by Thinkertech. Briefly, the raw signals were first adjusted to account for photo-bleaching by considering the overall trend before further analysis. To examine the response intensity of adenosine or ATP to light flickering stimulation, we obtained fluorescence change values (ΔF/F) by calculating (F − F0)/F0, where the baseline fluorescence signal (F0) was the average signal over a 300-s control time window. Peri-event plots were generated to display ΔF/F values.

#### Fiber photometry recording of optogenetic activation-evoked adenosine release

We utilized a three-color multi-channel fiber photometry device from Thinkertech to record optogenetic activation-evoked adenosine release. A single implanted ferrule was used to deliver a 580 nm stimulation laser (10 mW) coupled with a 470 nm blue LED light (30 μW). To investigate the impact of neuronal activation at different frequencies on adenosine release, we employed a 10/20/40 Hz square wave to modulate the excitation LED for photometry recording. During the ”off” period of the photometry LED, we administered laser pulses (power: 10 mW; pulse duration: 10%) for optogenetic stimulation. To measure the direct effect of neuronal activation on adenosine change, we recorded the optogenetically-evoked adenosine signal in freely moving mice.

### Polysomnographic recording and analysis

After at least one week of EEG/EMG implantation, the mice were housed individually in transparent barrels in a sound-proofed recording chamber with insulation and maintained on a 12-h light/12-h dark cycle with lights on at 7:00 am. They were provided with *ad libitum* access to food and water. The mice were acclimated to the recording cable for at least 3 days before starting the recording process. The cortical EEG and EMG signals were amplified, filtered with a high-pass filter above 0.5 Hz and band-pass filtered between 5-45 Hz, digitalized at a 1000 Hz resolution using a tethered data acquisition system (Medusa, Bio-Signal Technologies, China), and synchronized with the infrared video. The power of the 𝛿 (0.5–4 Hz), 𝜃 (5–8 Hz), and 𝛼 (9– 14 Hz) bands, as well as 𝛿 band ratio and EMG/activity signal, were calculated and scored (*42, 49*). For sleep behavior tests requiring light flickering treatment, light flickers of 20, 40, or 80 Hz at approximately 3000 lux were used. All light treatments were applied for 30 min just before the dark phase onset, and animals were kept awake by gentle handling if necessary to ensure light perception.

The sleep stages in the recordings were scored by AI-driven software, Lunion Stage, developed by LunionData (https://www.luniondata.com/en/lunion_stage). The AI engine of Lunion Stage employs a convolutional neural network, which is a type of deep learning model. This model extracts spectrogram features from the EEG channel and movement features from the EMG channel. After being trained on manually labeled recordings by domain experts, the model’s accuracy is assessed on recordings from diverse sources, including different devices and subjects, and achieves an average accuracy exceeding 98%. The EEG/EMG data collected were analyzed in 4-s epochs, and three stages, namely slow-wave sleep, rapid eye movement sleep and wakefulness, were automatically recognized based on their spectral and waveform properties(*50*). The scored results were examined and manual adjustments were made as needed. Based on the scored sleep stages, statistical analysis was performed on different vigilance states and groups.

### Histology and immunohistochemistry

To verify viral expression and optical fiber placement, mice were deeply anesthetized and immediately perfused with 0.1 M phosphate-buffered saline (PBS), followed by 4% paraformaldehyde (PFA). After being removed from the brain, the tissues were fixed overnight in 4% PFA and then dehydrated in a 30% sucrose solution. Brain samples were frozen in OCT compound (NEG-50, Thermo Scientific, USA), sectioned into 30-μm slices using a cryostat (HM525 NX, Thermo Scientific, USA), and stored at -20 °C until further processing.

Brain sections were prepared for immunostaining by permeabilization with PBST (0.3% Triton X-100 in PBS) for 30 min, blocking with 2% normal goat serum or normal donkey serum for 1 h, and then incubation with a primary antibody overnight at 4 °C. The brain sections were washed in PBS before being incubated with a secondary antibody. Finally, brain sections were washed in PBS and mounted with mounting media. The following primary or secondary antibodies were used in the current study at the indicated dilutions: chicken anti-mCherry (ab205402, abcam, 1:2000), rabbit anti-Cre (69050–3, Novagen, 1:3000), rabbit anti-GFAP (ab7260, abcam, 1:500), mouse anti-NeuN (ab77487, abcam, 1:500), rabbit anti-GABA (PA5-32241, Invitrogen,1:2000), guinea pig anti-vGluT2 (AB2251-1, abcam, 1:200), donkey anti-chicken Alexa Flour 488 (A78948, 1:1000), goat anti-guinea pig Alexa Fluor 647 (A-21450, Invitrogen, 1:1000), donkey anti-rabbit Alexa Fluor 594 (A-21207, Invitrogen, 1:1000), and goat anti-mouse Alexa Fluor 647 (A-21235, Invitrogen, 1:1000). The stained sections were visualized using either a confocal microscope (LSM900, Zeiss, Germany) or an epifluorescence microscope (DM6B, Leica, Germany).

### Targeted UHPLC-MS/MS

The mice were randomly assigned to one of three groups: the 40 Hz flickering visual stimulation group, the 30 min post-40 Hz flickering visual stimulation group, and the 240 min post-40 Hz flickering stimulation group. After flickering visual stimulation, the mice were sacrificed by rapid decapitation, and the hippocampus, striatum, mPFC, and visual cortex were promptly collected, frozen in liquid nitrogen, and stored at -80 °C until further analysis. To precipitate proteins from tissues, 200 μL of cold methanol: acetonitrile mixture (v:v = 1:1) was added, vortexed for 180 s, and the resulting supernatant was collected by centrifugation. The obtained supernatant was aliquoted into clean tubes, dried under N_2_ flow, and then redissolved in 100 μL of 1:1 (v:v) mixture of acetonitrile and water for subsequent UHPLC-MS/MS analysis.

A targeted assay was used to measure the levels of ATP, ADP, AMP, cAMP, IMP, homocysteine, SAH, SAM, methionine, inosine, and adenosine in V1 tissue. The analyses were conducted using a SHIMADZU CBM-30A Lite LC system together with an API 6500 Q-TRAP mass detector (AB SCIEX, Foster City, CA, USA), which was operated in ESI + mode. To separate the molecules, a Kinetex C18 100A column (100 ×2.1 mm, 2.6 μm) was utilized.

Standard solutions of ATP, ADP, AMP, cAMP, IMP, homocysteine, SAH, SAM, methionine, inosine, and adenosine (Sigma-Aldrich, purity >98%) were prepared at concentrations ranging from 1 to 1000 ng/mL and concentration calibration curves were established. Metabolite concentrations were determined using AB Sciex MultiQuant software (version 2.1, AB SCIEX, CA, USA).

### HPLC analysis of total tissue adenosine

The mice were randomly divided into four groups: normal light + vehicle, normal light + dipyridamole, 40 Hz flickering + vehicle, and 40 Hz flickering + dipyridamole. After the 30-min light flickering session, the mice were sacrificed by rapid decapitation, and the V1 area was dissected and stored at -80 °C until further analysis. To precipitate proteins from V1, a mixture of precooled adenosine inhibitors (500 μL of dipyridamole 1 mg/mL and 20 μL of EHNA 20 μg/mL) and acetonitrile in a 1:19 ratio (v:v) was added, and the sample was vortexed for 0.5 min and then centrifuged at 12,000 rpm for 15 min. The supernatant (100 µL) was injected into the HPLC system for analysis. Adenosine was separated by a Polaris 5 C18-A column (Agilent, 2.1 mm × 150 mm, 5 μm) at a temperature of 25 °C, with a detection wavelength of 258 nm. Isocratic elution was performed using a mobile phase consisting of 0.1% methanol, 0.1% (phase A), and 99.4% (phase B) at a flow rate of 1 mL/min. The mobile phase B was prepared with 25 mM sodium dihydrogen phosphate and adjusted to pH 7.0 using triethylamine.

### Real-time qPCR analysis

To obtain V1 tissue, mice were first anesthetized with isoflurane. Then, V1 tissues were microscopically dissected from the brain and quickly transferred to TRIzol reagent (Invitrogen, CA, USA). RNA quantity and quality were assessed using a NanoDrop spectrophotometer. The reverse transcription process was carried out using a PrimeScriptTM RT reagent kit (Vazyme,

R323, Nanjing, China). The primers used for qRT-PCR included ENT1 (5’- CAGCCTCAGGACAGGTATAAGG-3’ and 5’-GTTTGTGAAATACTTGGTTGCGG-3’), ENT2 (5’- TCATTACCGCCATCCCGTACT-3’ and 5’- CCCAGTTGTTGAAGTTGAAAGTG-3’), ADA (5’-ACCCGCATTCAACAAACCCA-3’ and 5’-AGGGCGATGCCTCTCTTCT-3’), and ADK (5’-AGAGTCGGTATTGAAAGTGGCT-3’ and 5’-CTAAAGCAACTCACCGTCTCAT-3’). The expression levels of these genes were normalized based on the housekeeping gene β-actin. The SYBR Green fluorescence system was used for quantitative mRNA analysis with a StepOnePlus Real-Time PCR System (Life Technologies, USA), following the manufacturer’s instructions. Then, the mRNA expression level was determined using the ΔΔCt method, and β-actin was used as the calibrator gene. Target gene expression was determined as relative expression levels and each sample was analyzed at least in triplicates.

### Clinical study on the effect of 40 Hz flicker on children with insomnia symptoms

#### Participant recruitment

The study protocol (#2022-130-K-99-01) was approved by the Ethics Committees of the School of Optometry and Ophthalmology and Eye Hospital of Wenzhou Medical University, and the study was carried out following the Declaration of Helsinki. This study was conducted between February 2022 and May 2023 at the Department of Pediatrics, Second Affiliated Hospital, and Yuying Children’s Hospital of Wenzhou Medical University. Demographic data were collected from pediatricians, and any identifying information was eliminated to ensure anonymity. Data analysis was conducted by investigators blind to patients’ data and study design and then interpreted by all authors. Potential participants and their parents were provided with detailed information regarding the purpose and procedures of the study, and they were required to sign an informed consent form before participating. The inclusion criteria were as follows: (1) children aged 4 to 16 years, (2) a diagnosis of insomnia for the first time according to the International Classification of Sleep Disorders, Third Edition (ICSD-III) without any prior treatment, and (3) signed informed consent forms from both patients and their parents. The study’s exclusion criteria were: (1) a history of neurological, psychiatric, or sleep disorders, (2) caffeine abuse, or the use of psychotropic medications within the past 12 months.

#### Procedures

The study followed a self-controlled design, and polysomnographic (PSG) recordings were performed for three consecutive nights on all enrolled patients. On the first day, baseline sleep parameters were measured. On day 2, patients underwent a 30-min session of 40 Hz light flickering using a flickering glass device (Hangzhou Chongzheng Medical Co., Ltd) from 8:30 to 9:00 pm, and sleep parameters were recorded for the entire night. Demographic data, such as the participant’s name, gender, weight (kg), height (cm), and calculated Body Mass Index (BMI), were collected. Before the study began, all patients underwent extensive history recording, including presenting complaints and history of presenting complaints, and were given half a day to acclimate to the ward environment. Whole-night sleep recordings were obtained using PSG equipment (Embla s4500, USA). Airflow was evaluated using a nasal pressure and/or oronasal thermistor sensor, while heart rate and arterial blood oxygen saturation levels were monitored using a pulse oximeter (Nonin Medical Inc.) during night-time sleep. These parameters were obtained simultaneously and continuously throughout the night. Sleep stages were classified based on the 2012 Academy of Sleep Medicine (AASM) manual guidelines(*48*).

#### Outcomes

The primary objective was to evaluate the effects of 40 Hz flickering on sleep onset by measuring sleep onset latency (SOL, minutes from light off to sleep onset) with polysomnography (PSG) and comparing it to baseline. The secondary outcomes were to assess improvement in sleep maintenance parameters such as total sleep time (TST, sleep period time minus the duration of intra-sleep awakenings), sleep efficiency (SE, total sleep time/total time in bed), wake time after sleep onset (WASO, minutes of wake after sleep onset) and arousal frequency (AF, the number of nightly awakenings) compared to baseline. In addition to primary and secondary endpoints, this study also evaluated other sleep parameters including REM sleep onset latency (REM SOL, the time in minutes between sleep onset and the first epoch of REM sleep), percentage of light sleep (N1+N2) and percentage of deep sleep (N3).

#### Statistical Analysis

GraphPad Prism 9.0 software (GraphPad Software) was utilized for statistical analysis. Data normality was checked using the Kolmogorov-Smirnov test. If the data were normally distributed, the mean ± SD was used to express it, and a one-way repeated-measures analysis followed by Tukey’s multiple comparison test was applied. Data that did not meet a normal distribution were reported as median (P25-P75) and analyzed with the Friedman test, followed by Dunn’s multiple comparison test. P-value < 0.05 was considered statistically significant.

## Declarations

### Ethics approval and consent to participate

All animal experiments and procedures involving animals were approved by the Animal Care and Use Committee of Wenzhou Medical University (NO. wydw2021-0563). The clinical study was approved by the Institutional Ethics Board Committee of the Wenzhou Medical University Eye Hospital on human experimentation before study initiation and was carried out in compliance with the World Medical Association Declaration of Helsinki (NO. 2022-130-K-99- 01).

### Consent for publication

All authors have approved the contents of this manuscript and provided consent for publication.

### Availability of data and materials

Not applicable.

### Author contributions

Conceptualization: J.-F.C., Y.H., X.Z., and T.X.; Methodology: Y.H., X.Z., and T.X.; Formal analysis: Y.H., Z.X., T.X., X.H.C., Y.L., Y.Z., X.X., H.S., Z.Y., ZW.L., T.F., and S.Z.; Investigation: J.-F.C., Y.H., X.Z., T.X., W.G., and Z.L. Resources: Y.L., Z.W., Z.H., and J.Q.; Writing - Original Draft: J.-F.C., Y.H., X.Z., and T.X.; Writing - Review & Editing: W.G., J.-F.C., Y.H., R.A.C, X.Z., and T.X.; Visualization: Y.H., X.Z., T.X., and Y.L.; Supervision: J.-F.C.; Project administration: J.-F.C. and J.Q.; Funding acquisition: J.- F.C., X.Z., W.G., and Z.L.

## Data Availability

All data produced in the present study are available upon reasonable request to the authors

## Acknowledgments

We thank Drs. Shengtao Hou, Jianhong Zhu, Yuanguo Zhou, Weihong Song, and Libin Huang for critical readings, discussion and suggestions to improve the manuscript.

