## Supplemental Data1 for "40 Hz Light Flickering Promotes Sleep through Cortical Adenosine Signaling": Supplement figure.pdf

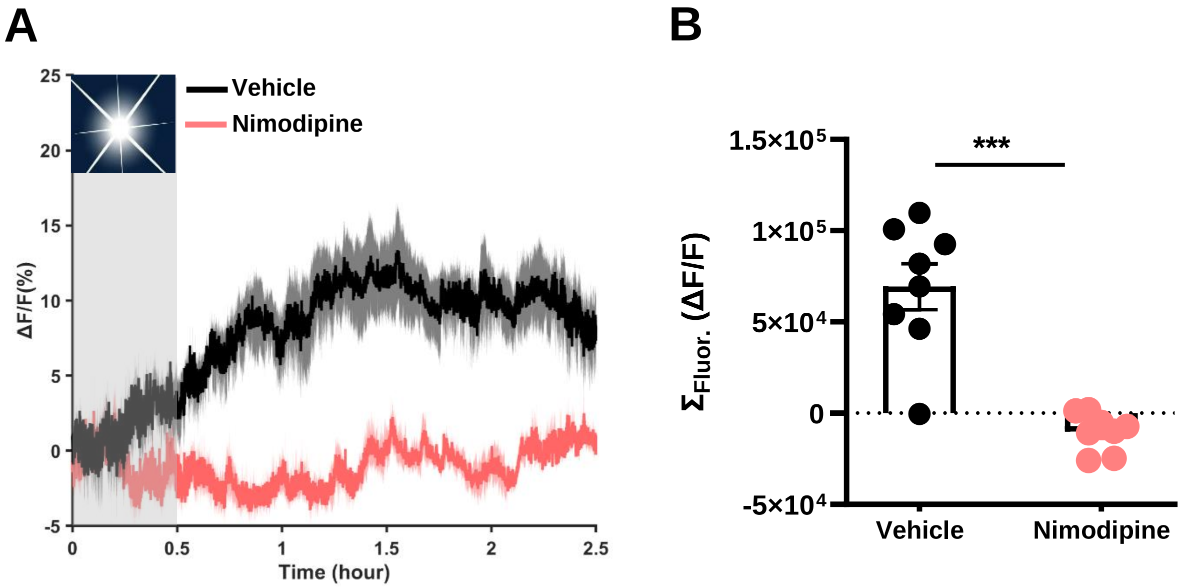


**FigureS1. Activity-dependent Ado release is blocked by inhibitors of L-type voltage gated calcium channels (VGCCs)**

**A.** Pretreatment with the L-type VGCCs antagonist nimodipine (10 mg/kg, i.p.) abolished the extracellular adenosine induction by 40 Hz flickering. **B.** The quantification of visual flickering-evoked Ado signals. ***$P < 0.001$, Student's t-test. The data are presented as Mean±SEM.

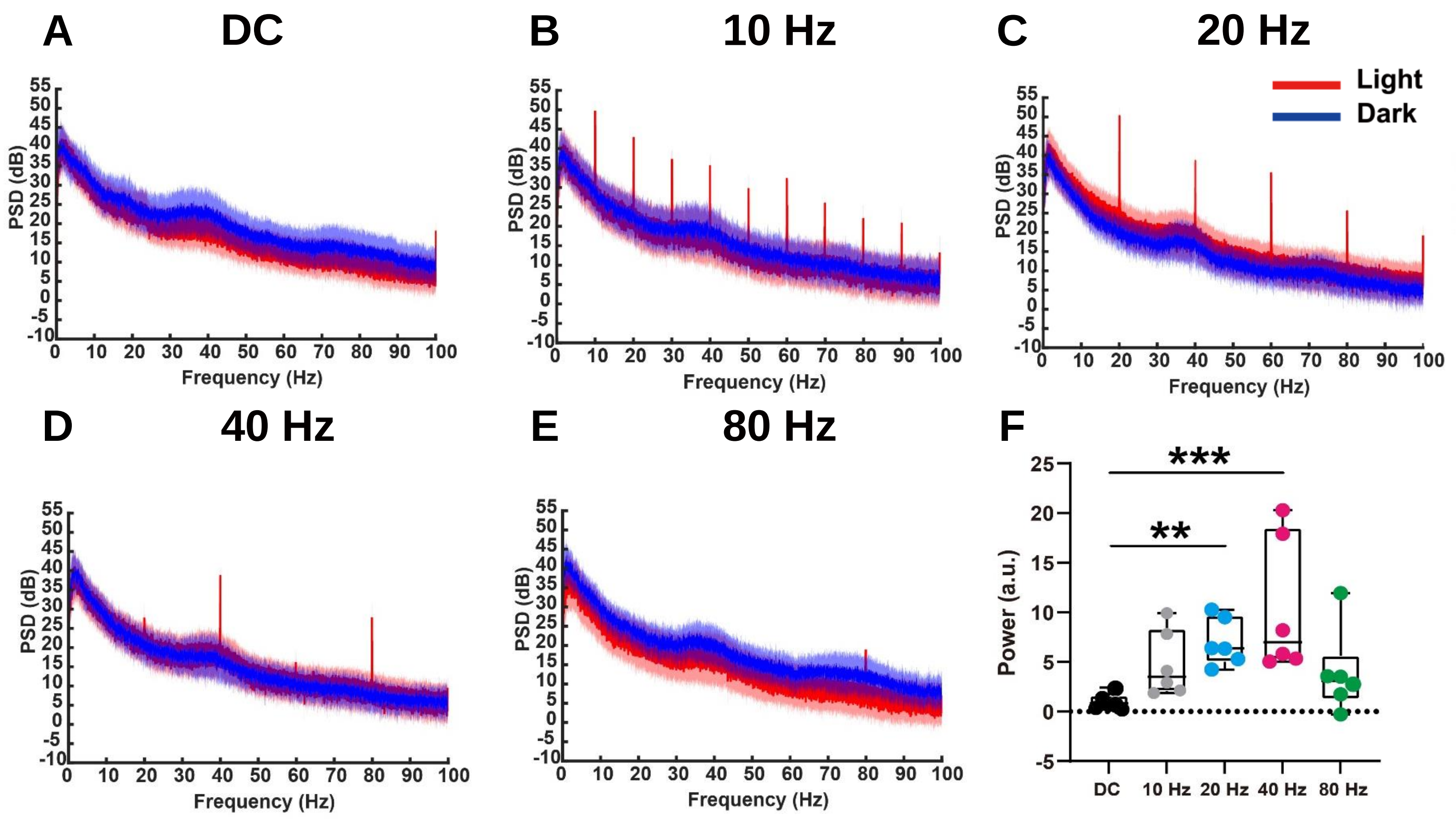


**FigureS2. Each frequency of light flickering elicited a specific LFP oscillation at that frequency, as well as at its higher harmonic frequencies.**

The mean power spectra ± SEM of DC (**A**), 10 Hz (**B**), 20 Hz (**C**), 40 Hz (**D**), and 80 Hz (**E**) flickering in V1 during no-stimulation (5 min, blue line) and stimulation (30 min, red line), overlayed, n = 6/group.

**F.** The boxplot showed the quantified and normalized value of power (same frequency as the flickering) during flickering (n = 6/group). The skewed data were analyzed using the Friedman test for repeated measures, followed by Dunn's multiple comparisons test. ***P < 0.001, **P < 0.01. Data are presented as Mean±SEM

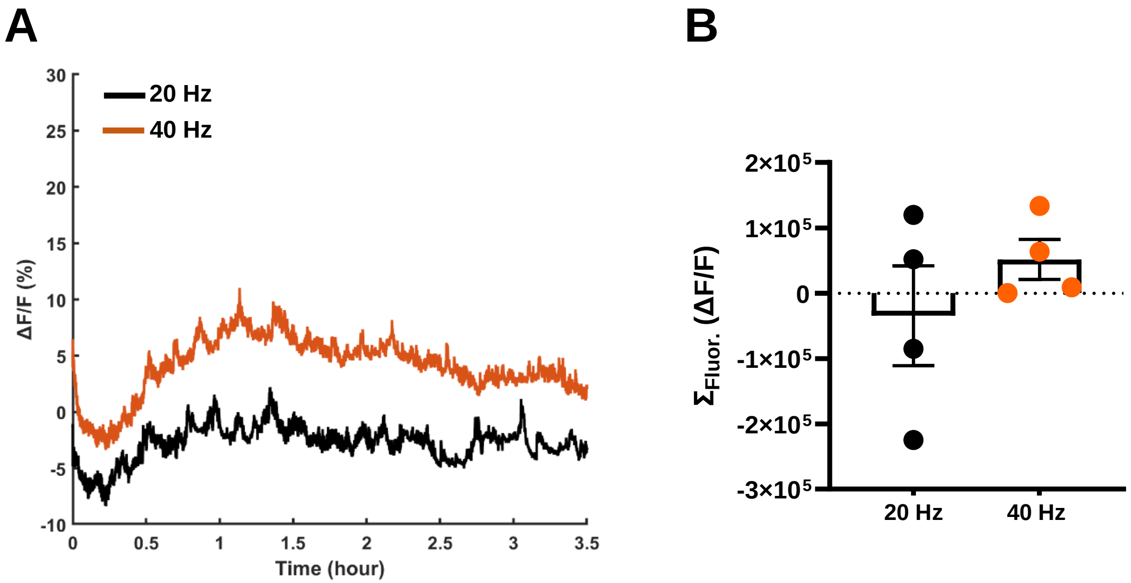


**FigureS3. Frequency-dependent of light flickering on the extracellular adenosine levels in the basal forebrain**

**A.** Extracellular adenosine levels increased more in response to 40 Hz light flickering compared to 20 Hz and light flickering (The 40 Hz light flickering data has been shown in the figure2I ) (*n* = 4/group). **B.** The quantification of visual flickering-evoked Ado signals. The data are presented as Mean±SEM.

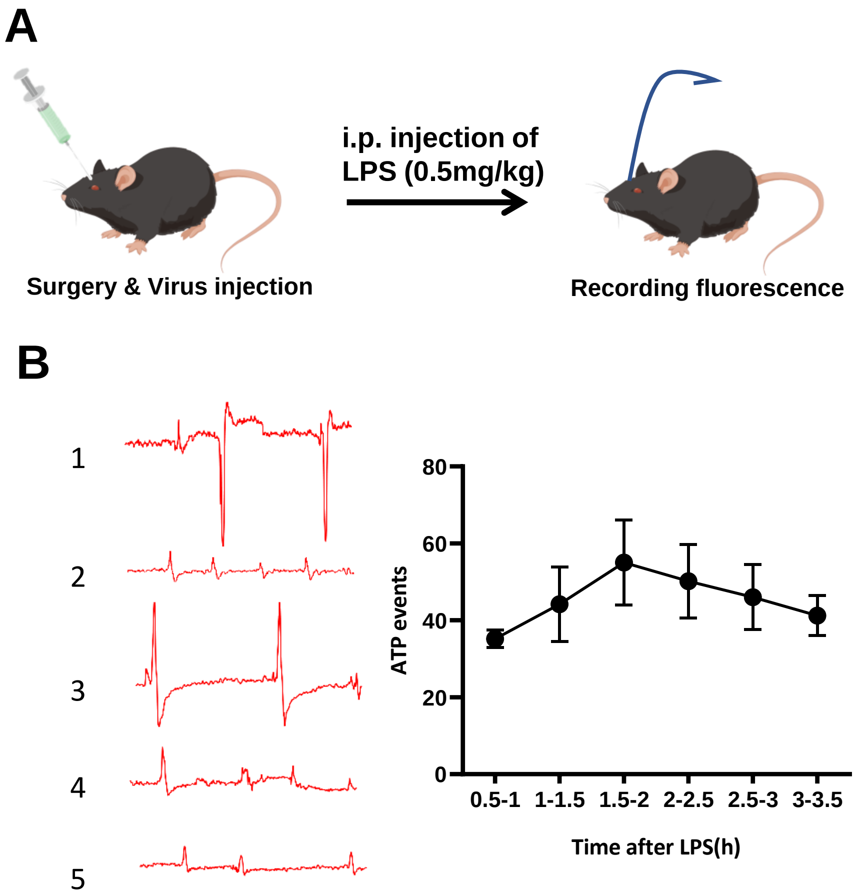


**FigureS4. GRAB$_{ATP1.0}$ reveals localized ATP-release events in the visual cortex following lipopolysaccharides (LPS)-induced systemic inflammation**

**A.** Schematic diagram depicting the experimental protocol in which an AAV encoding CRE under the control of the promoter hSyn is injected into the GRAB$_{ATP1.0}$-KI mice in the visual cortex, followed by using fiber photometry to assess fluorescence signals after an intraperitoneal injection of LPS.

**B.** left: Schematic representation of ATP levels in each mouse following LPS injection; right: Measurement of extracellular ATP levels after 0.5 mg/kg LPS treatment in GRAB$_{ATP1.0}$-KI mice. Data are presented as Mean±SEM.
